# Health, socioeconomic and genetic predictors of COVID-19 vaccination uptake: a nationwide machine-learning study

**DOI:** 10.1101/2022.11.11.22282213

**Authors:** Tuomo Hartonen, Bradley Jermy, Hanna Sõnajalg, Pekka Vartiainen, Kristi Krebs, Andrius Vabalas, FinnGen, Estonian Biobank Research Team, Tuija Leino, Hanna Nohynek, Jonas Sivelä, Reedik Mägi, Mark Daly, Hanna M. Ollila, Lili Milani, Markus Perola, Samuli Ripatti, Andrea Ganna

**Affiliations:** Institute for Molecular Medicine Finland, FIMM, HiLIFE, University of Helsinki, Helsinki, Finland; Estonian Genome Centre, Institute of Genomics, University of Tartu, Tartu, Estonia; The Finnish Institute for Health and Welfare, Helsinki, Finland; Broad Institute of MIT and Harvard, Cambridge, MA, USA; Massachusetts General Hospital, Cambridge, MA, USA; Harvard Medical School, Cambridge, MA, USA; Center of Genomic Medicine, Harvard Medical School, Boston MA, USA; Anesthesia, Critical Care, and Pain Medicine, Massachusetts General Hospital and Harvard Medical School, Boston, MA, USA; Department of Public Health, University of Helsinki, Helsinki, Finland

**Author notes:** These authors contributed equally (Tuomo Hartonen, Bradley Jermy). Estonian Biobank research team: Andres Metspalu, Tõnu Esko, Reedik Mägi, Mari Nelis and Georgi Hudjashov.

## Abstract

Reduced participation in COVID-19 vaccination programs is a key societal concern. Understanding factors associated with vaccination uptake can help in planning effective immunization programs. We considered 2,890 health, socioeconomic, familial, and demographic factors measured on the entire Finnish population aged 30 to 80 (N=3,192,505) and genome-wide information for a subset of 273,765 individuals. Risk factors were further classified into 12 thematic categories and a machine learning model was trained for each category. The main outcome was uptaking the first COVID-19 vaccination dose by 31.10.2021, which has occurred for 90.3% of the individuals.

The strongest predictor category was labor income in 2019 (AUC evaluated in a separate test set = 0.710, 95% CI: 0.708-0.712), while drug purchase history, including 376 drug classes, achieved a similar prediction performance (AUC = 0.706, 95% CI: 0.704-0.708). Higher relative risks of being unvaccinated were observed for some mental health diagnoses (e.g. dissocial personality disorder, OR=1.26, 95% CI : 1.24-1.27) and when considering vaccination status of first-degree relatives (OR=1.31, 95% CI:1.31-1.32 for unvaccinated mothers)

We derived a prediction model for vaccination uptake by combining all the predictors and achieved good discrimination (AUC = 0.801, 95% CI: 0.799-0.803). The 1% of individuals with the highest risk of not vaccinating according to the model predictions had an average observed vaccination rate of only 18.8%.

We identified 8 genetic loci associated with vaccination uptake and derived a polygenic score, which was a weak predictor of vaccination status in an independent subset (AUC=0.612, 95% CI: 0.601-0.623). Genetic effects were replicated in an additional 145,615 individuals from Estonia (genetic correlation=0.80, 95% CI: 0.66-0.95) and, similarly to data from Finland, correlated with mental health and propensity to participate in scientific studies. Individuals at higher genetic risk for severe COVID-19 were less likely to get vaccinated (OR=1.03, 95% CI: 1.02-1.05).

Our results, while highlighting the importance of harmonized nationwide information, not limited to health, suggest that individuals at higher risk of suffering the worst consequences of COVID-19 are also those less likely to uptake COVID-19 vaccination. The results can support evidence-informed actions for COVID-19 and other areas of national immunization programs.

## Introduction

In the face of the worldwide COVID-19 pandemic, safe and effective vaccines were developed and approved for use in record-breaking time [1]. However, across high-income countries, somewhere between 5% and 30% of the population has not received any dose of COVID-19 vaccine, and higher proportions of unvaccinated were observed in low-income countries [2]. In Finland, 23.5% of the population has not received any dose of COVID-19 vaccine by the end of October 2021, in line with several other European countries. In the case of COVID-19, a comprehensive and rapid vaccination of the population is key to reducing disease severity (vaccination effectiveness, VE, against death 99.0% [3]), alleviating the healthcare burden (VE against hospitalization 97.2% [3]) and reducing the spread of infection [4]. Refusal, postponement, or inability to participate in the vaccination program is therefore a key societal concern. Being able to identify individual factors impacting vaccination uptake can help policymakers to design more effective targeted interventions for future immunization programs.

Previous studies aiming to identify factors underlying the intention to take a COVID-19 vaccination were mostly based on surveys [5,6,7,8,9,10,11]. These studies have identified factors such as trust and knowledge of the COVID-19 vaccines, recommendations by healthcare professionals, as well as beliefs about the disease severity and convenience of vaccination as important predictors of intention to take COVID-19 vaccination. This is in line with previous studies about vaccine hesitancy [12,13]. While offering important information about self-perceived reasons for vaccine hesitancy, studies based on survey data have several limitations. First, surveys are typically limited to a few thousand individuals, limiting the power to identify individuals at risk. Second, survey participants are often not representative of the general population and factors associated with vaccine hesitancy, such as socioeconomic status or education level, are also associated with participation in scientific studies [14]. Thus, individuals more likely to not participate in a vaccination program are also more likely to be under-represented in surveys. Third, surveys are affected by reporting bias, either voluntary or involuntary, and can collect only a limited set of information limiting the power of epidemiological and machine learning analyses.

To address these limitations and to provide new insights, we used a comprehensive collection of nationwide registers covering detailed health, socioeconomic, familial and demographic information to map potential predictors of COVID-19 vaccination uptake across the entire Finnish population (5.5 million individuals). We compared 2,890 predictors measured before 31.12.2019 and uptake of the first dose of COVID-19 vaccine between 27.12.2020 and 31.10.2021. We used machine learning methods to quantify the importance of 12 different predictor categories (e.g. disease history, medication purchases, education level, see **Figure 1a**) and their overlap. Finally, we combined these categories to derive a prediction model of COVID-19 vaccination status.

**Figure 1.**
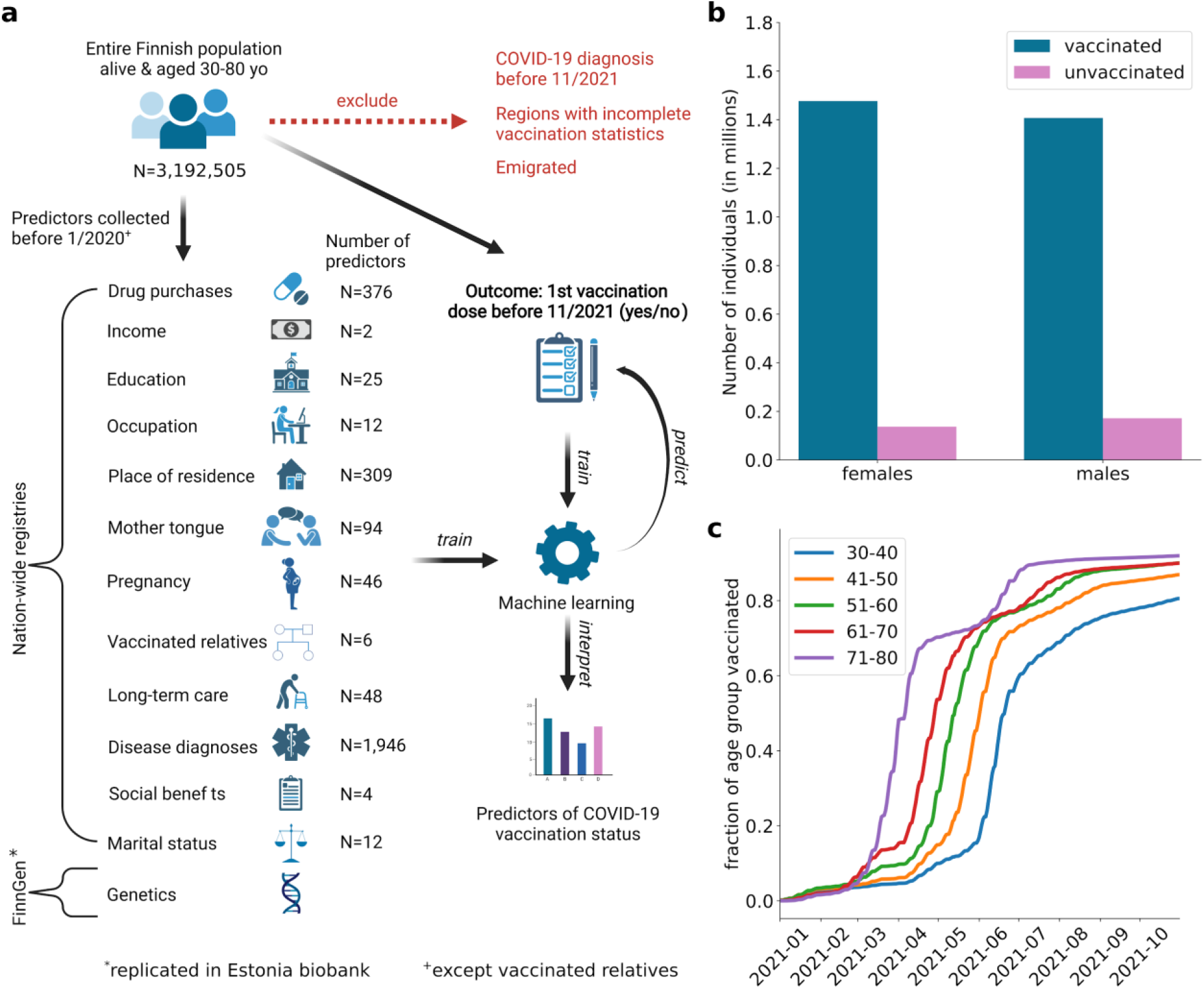
**a)** Schematic outline of the study. COVID-19 vaccination uptake (at least one vaccination dose) at the end of October 2021 was extracted from the Finnish Vaccination Register for each individual aged 30-80 years and living in Finland. A comprehensive collection of potential predictors was extracted (at the end of 2019, except for vaccination status of relatives for which data up to end of October 2021 was used) from nation-wide registries, totalling 2,890 potential predictors across 12 manually defined predictor categories. Genetics of Covid-19 vaccination uptake was studied in a subsample of individuals of the total study population (FinnGen participants) and replicated in Estonia Biobank. Machine learning was then used to identify predictors and predictor categories that best predict vaccination uptake in the test set. **b)** Total number of vaccinated (blue, at least one vaccination dose) and unvaccinated (purple) females and males in the study population at the end of October 2021. **c)** Cumulative fraction of different age groups in the study population (blue: 30-40 year olds, orange: 41-50 year olds, green: 51-60 year olds, red: 61-70 year olds, violet: 71-80 year olds) who have received 1st dose of COVID-19 vaccine as function of time during the follow-up period.

Previous studies have shown a genetic liability and identified individual genetic factors that impact COVID-19 severity and susceptibility [15]. Across 273,765 individuals (with replication in additional 145,615 individuals from Estonia), we evaluated if genetic information could predict COVID-19 vaccination uptake, if there is a genetic overlap with health and behavioral traits that were not available nationwide and if individuals with higher genetic risk for COVID-19 were more or less likely to be vaccinated.

Understanding the predictors of vaccination uptake is an important step toward a more sustainable public health response. This study establishes a framework for using machine learning and statistical genetics methods to identify individuals that are less likely to participate in COVID-19 vaccination programs.

## Results

### Comprehensive nationwide information to identify predictors of COVID-19 vaccination uptake

The FinRegistry project (https://www.finregistry.fi/) combines and harmonizes data from 18 Finnish nationwide registers into a comprehensive dataset for epidemiological and machine learning analyses. Briefly, these registers cover disease diagnoses from primary, secondary and tertiary care, medication purchases, welfare benefits, multi-generational familial relationships, socio-economic and demographic information for at least 10 years, with some registers dating back to the 1970’s (**Figure 1** for study overview, **Data and Methods**). One of these registers, the Finnish Vaccination register, contains records of all COVID-19 vaccination doses administered in Finland.

We manually divided the vast amount of information, in total 2,890 potential predictors, into 12 consistent categories for easier interpretation of the results. Predictors were available before 31^st^ December 2019 (i.e. before the start of the COVID-19 pandemic, except for the vaccination status of relatives for which vaccination records until 31st October 2021 were used) for all individual residents of Finland alive on the 31st December 2020. We considered only individuals between 30 and 80 years old and excluded 6.1% of the study population who had emigrated and a further 1.9% with a reported positive COVID-19 test by the 31^st^ October 2021. We further excluded 0.1% of the remaining study population living in Askola, a municipality with incomplete vaccination records (see **Extended Data Figure 1**). We chose the age range 30-80 because by 31^st^ October 2021 everyone in this age range had been eligible for a first dose of COVID-19 for at least 4 months (**Figure 1c**). In total we included 3,192,505 individuals (50,5% females), of which 136,947 females (8.5%) and 171,647 males (10.9%) (**Figure 1b**) were unvaccinated. Younger individuals were eligible for vaccination later and had a lower vaccination rate by the end of the study period (**Figure 1c**). Thus, age was used as covariate in all the presented analyses. Genetic information from the FinnGen study [16] was available for a subset of 273,765 individuals fulfilling similar inclusion criteria, of which 93% had received their first dose of a COVID-19 vaccination by 31.10.2021. Details of data preprocessing are reported in the **Data and Methods** section.

### Income in 2019 and drug purchase history were the strongest predictors of COVID-19 vaccination uptake

We studied the importance of the 12 categories of predictors in predicting COVID-19 vaccination uptake using machine learning models (XGBoost [17]) trained separately for each category in a randomly sampled 80% of the study population and evaluated on the remaining 20%. Each model also included age and sex as predictors, representing the baseline model. Income (area under receiver-operator characteristics curve (AUC) = 0.710, 95% CI: 0.708-0.712) and history of previous drug purchases, including 376 drug classes, (AUC = 0.706, 95% CI: 0.704-0.708) were the most predictive categories (**Figure 2a, Supplementary Table 1**). All but one of the categories, long-term care, performed better than the simple baseline model including only age and sex (AUC=0.612, 95% CI: 0.610-0.614; **Figure 2a**).

**Figure 2.**
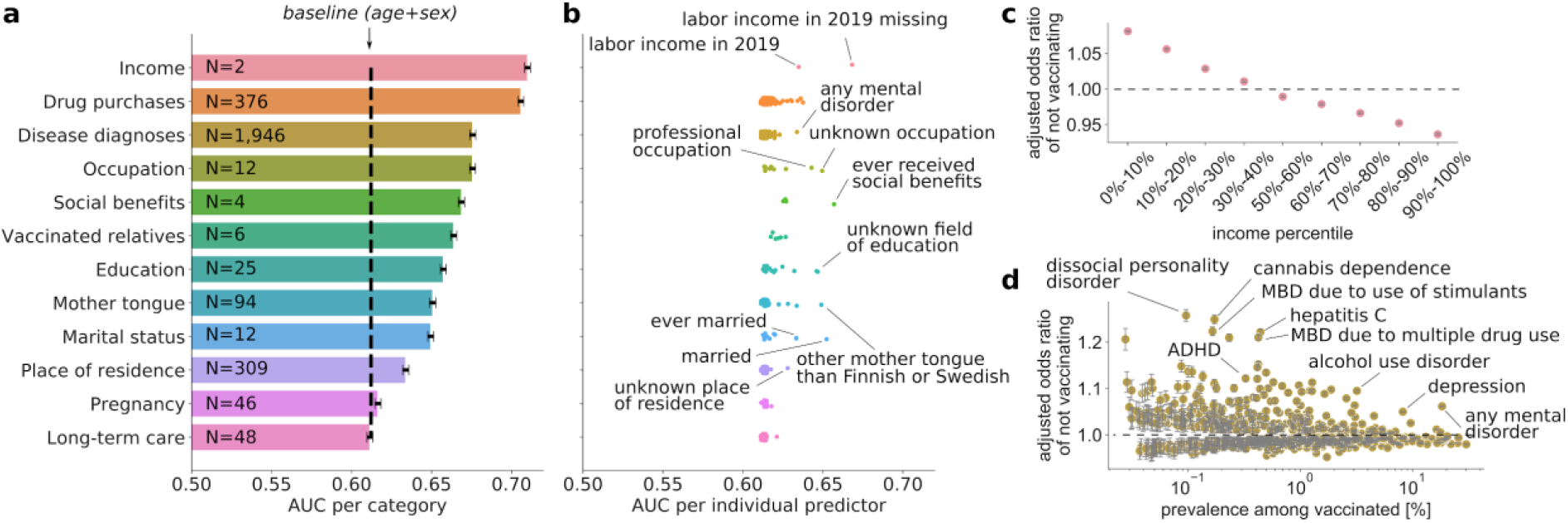
**a)** Area under receiver-operator characteristics curve (AUC) for XGBoost classifiers trained using predictors from different predictor categories (each model also includes the baseline predictors age and sex). Error bars show 95% confidence intervals computed using bootstrapping. Number of predictors within each category is indicated on top of the corresponding bar. The black vertical dashed line indicates the performance of an XGBoost model allowed to use only age and sex as predictors. Most of the predictor categories perform better than this baseline model, with Income and Drug purchases being the most predictive categories. **b)** AUC from Lasso classifiers trained separately for each of the individual predictors (models also include the baseline predictors age and sex), grouped by the categories. Some of the interesting highly predictive predictors have been highlighted (for a fully annotated list of AUCs of individual predictors, see **Supplementary Table 2**). **c)** Association between labor income in 2019 and COVID-19 vaccination uptake. Odds ratio from a logistic regression model using income percentile bins as predictors and adjusting for age and sex. The 40%-50% percentile bin was used as a reference category. Error bars indicate 95% confidence intervals for odds ratios computed using bootstrapping. **d)** Associations between previous disease diagnoses and COVID-19 vaccination status. Odds ratio from a logistic regression model using a binary disease indicator as predictor and adjusting for age and sex. Some of the interesting predictors are highlighted. Predictors with multiple hypothesis testing-adjusted p-value > 0.01 (Benjamini-Hochberg method), and prevalence among vaccinated <1000 are not shown. Error bars indicate 95% confidence intervals for odds ratios computed using bootstrapping. For a fully annotated list of ORs of individual predictors, see **Supplementary Table 3**.

Next, we studied the classification performance of individual predictors within each category by training individual Lasso models for each of the 2,890 predictors, including the baseline variables age and sex (**Figure 2b, Supplementary Table 2**; see **Data and Methods** for details). To provide interpretable effect sizes, we also performed logistic regression (without penalization) for each of the predictors, including age and sex as covariates, and calculated odds ratios (OR) of not vaccinating against COVID-19 (**Extended Data Figure 2, Supplementary Table 3**; see **Data and Methods**). Reference levels for the predictors used in the logistic regression analysis are listed in **Supplementary Table 8**. Not having income from labor in 2019 was the most predictive individual predictor (AUC=0.668, 95% CI: 0.666-0.671, OR=1.35, 95% CI: 1.35-1.35). Among individuals with labor income, those in the lowest income decile had a significantly higher chance of not uptaking vaccination compared to individuals in the 40%-50% income decile bin (OR=1.08, 95% CI: 1.08-1.09; **Figure 2c**). Overall we observed a linear relationship between income and COVID-19 vaccination uptake. Other socio-demographic variables such as speaking another mother tongue than Finnish or Swedish were both strong predictors and conferred an elevated relative risk of not vaccinating (AUC=0.649, 95% CI: 0.647-0.651; OR=1.27, 95% CI: 1.27-1.27).

We examined individual disease diagnoses (**Figure 2d**) and drug purchases to identify possible disease groups associated with vaccination uptake (**Supplementary Table 3**). The highest odds ratios of not vaccinating were observed for diagnoses of substance abuse, such as stimulants (OR=1.22, 95% CI : 1.21 - 1.23) and cannabinoids (OR=1.25, 95% CI:
1.24 - 1.26), and also for hepatitis C diagnosis (OR=1.22, 95% CI 1.21 - 1.23) which is strongly associated with intravenous drug usage. Other mental health conditions, particularly those associated with psychotic- or delusion-type symptoms, showed large relative risks (e.g. OR of dissocial personality disorder = 1.24, 95% CI: 1.23 - 1.26, OR of schizoid personality disorder = 1.14, 95% CI: 1.13 - 1.15).

While drug purchase history was the second strongest predictor category, no single medication alone was a strong predictor, suggesting that the combined history of different drug purchases is largely responsible for the predictiveness of the category. However, several of the most predictive drugs associated with not vaccinating were those used in the treatment of psychosis-associated disorders, such as phenothiazines (OR 1.07, 1.07 - 1.07) and novel/atypical antipsychotics (OR 1.07, 1.07-1.07). ADHD medications had the highest OR among the individual drugs (1.08, 95% CI 1.07 - 1.09). Memantine, a medication used to treat symptoms of cognitive impairment such as Alzheimer’s disease, was associated with a higher non-vaccination rate (OR 1.04 and 95% CI 1.03-1.05).

Because of comprehensive information on multi-generational familial relationships, we could study how the vaccination status of a close relative impacts the chances of taking up vaccination (**Extended Data Figure 3**). We only considered individuals with relatives in the study population (see **Supplementary Table 3, Data and Methods**).

We found that having an unvaccinated mother increases the risk of not being vaccinated (OR=1.31, 95% CI:1.31-1.32). The risk of not vaccinating was smaller when having an unvaccinated father (OR=1.23, 95% CI: 1.22-1.23) or when having any unvaccinated siblings (OR=1.17, 95% CI: 1.16-1.17).

We performed a sensitivity analysis to account for individuals not eligible for vaccination due to non-reported emigration outside Finland. To capture unreported emigration, we excluded all individuals with no data entries in 2019 (4.0%; see **Data and Methods**). Overall, we did not observe differences in predictive performance for most individual predictors as measured by AUC (**Extended Data Figure 4a**). However, we observed significant deflation in the ORs of several rare mother tongues (**Extended Data Figure 4b**). The OR for speaking another mother tongue than Finnish or Swedish decreased from 1.27 to 1.15.

### A prediction model for COVID-19 vaccination uptake

Combining all the registry-based predictors into a single XGBoost model resulted in good discrimination (AUC = 0.801, 95% CI: 0.799-0.803 in the test set) but modest calibration. However, we recalibrated the model using the method from ref. [18] obtaining a good calibration (**Extended Data Figure 5**). In the test set, the top 1% of individuals with the highest predicted probability of not uptaking vaccination (N=6,385) had an observed vaccination rate of only 18.8% vs 90.3% when considering everyone in the test set (**Figure 3a**). The XGBoost classifier outperformed a Lasso classifier trained using the same full set of predictors (AUC=0.778, 95% CI: 0.776-0.780).

**Figure 3.**
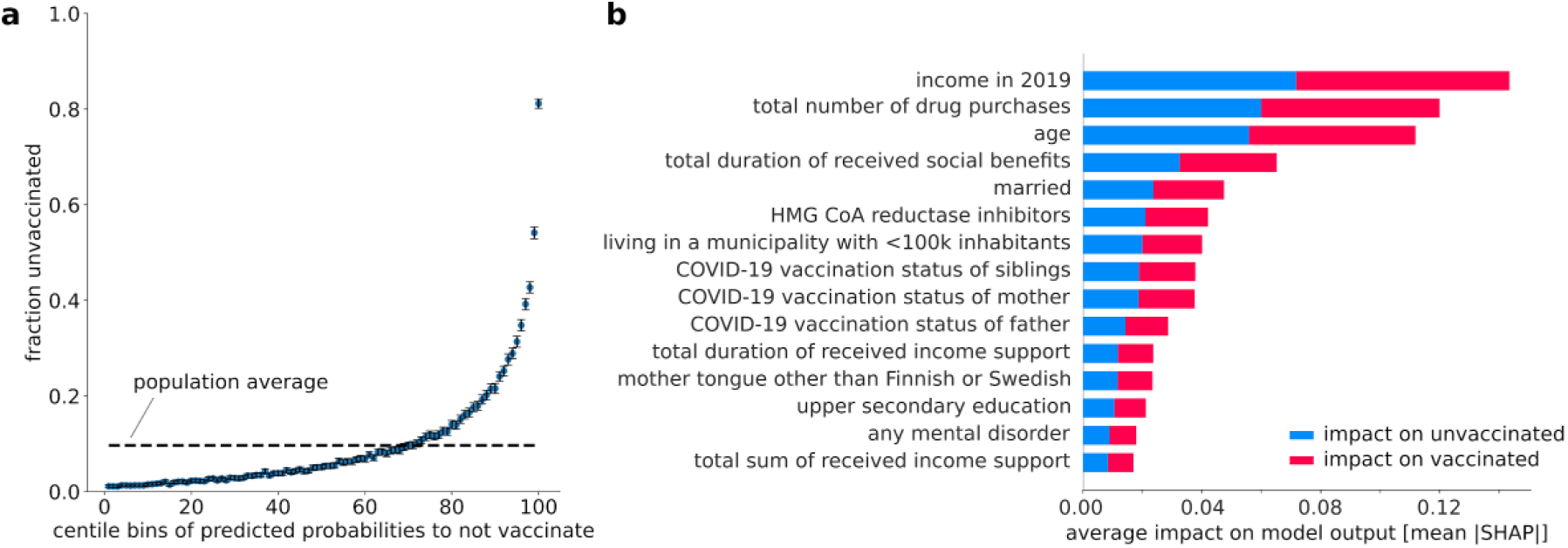
**a)** Fractions of unvaccinated individuals in the test set as a function of centile bins of predicted probabilities to not vaccinate from the full XGBoost model. The 99th centile bin comprises 6,385 individuals that have only an 18.8% (95% CI: 17.9%-19.8%) chance of vaccinating. The error bars indicate 95% confidence intervals computed using bootstrapping. The black dashed line indicates the average fraction of unvaccinated individuals in the study population. **b)** Mean absolute Shapley values [16] computed for all individual predictors used in the full XGBoost model. Higher values indicate higher average impact of the predictor in pushing the model output towards unvaccinated (blue), and vaccinated (red). The top 20 most important predictors are shown for clarity.

We analyzed the importance of each predictor in the combined XGBoost model by computing the mean absolute Shapley values of the predictors [19]. A handful of predictors have a strong contribution to the model, with income, the total number of drug purchases, age, total duration of received social benefits, and being married among the most important predictors (**Figure 3b**). Interestingly, income is a stronger predictor of COVID-19 vaccination status than age.

### Different predictor categories share similar information on COVID-19 vaccination uptake

Some of the predictor categories included in this study are often considered to capture separate information. For example, information on drug purchases should mainly capture health, while income and job profession should represent important socio-economic factors. To study how much independent information each predictor category contains, we considered all possible combinations of predictor categories, and trained a separate Lasso classifier model for each of the 4,097 combinations. The rationale for this experiment is that, by testing each possible combination of predictors categories, we can quantify information relevant to COVID-19 vaccination prediction that is unique to single categories *vs* what is shared across categories.

**Figure 4a** shows the drop in AUC when removing each predictor category separately from the combined model. As expected, classification performance decreases the most when removing the drug purchases history category, leading to a drop in AUC of 1.3%. However, this decrease is substantially lower than the AUC improvement that this category contributes on top of age and sex (15.3%), indicating that much of the predictive information from this category, is captured by other categories present in the combined model.

**Figure 4.**
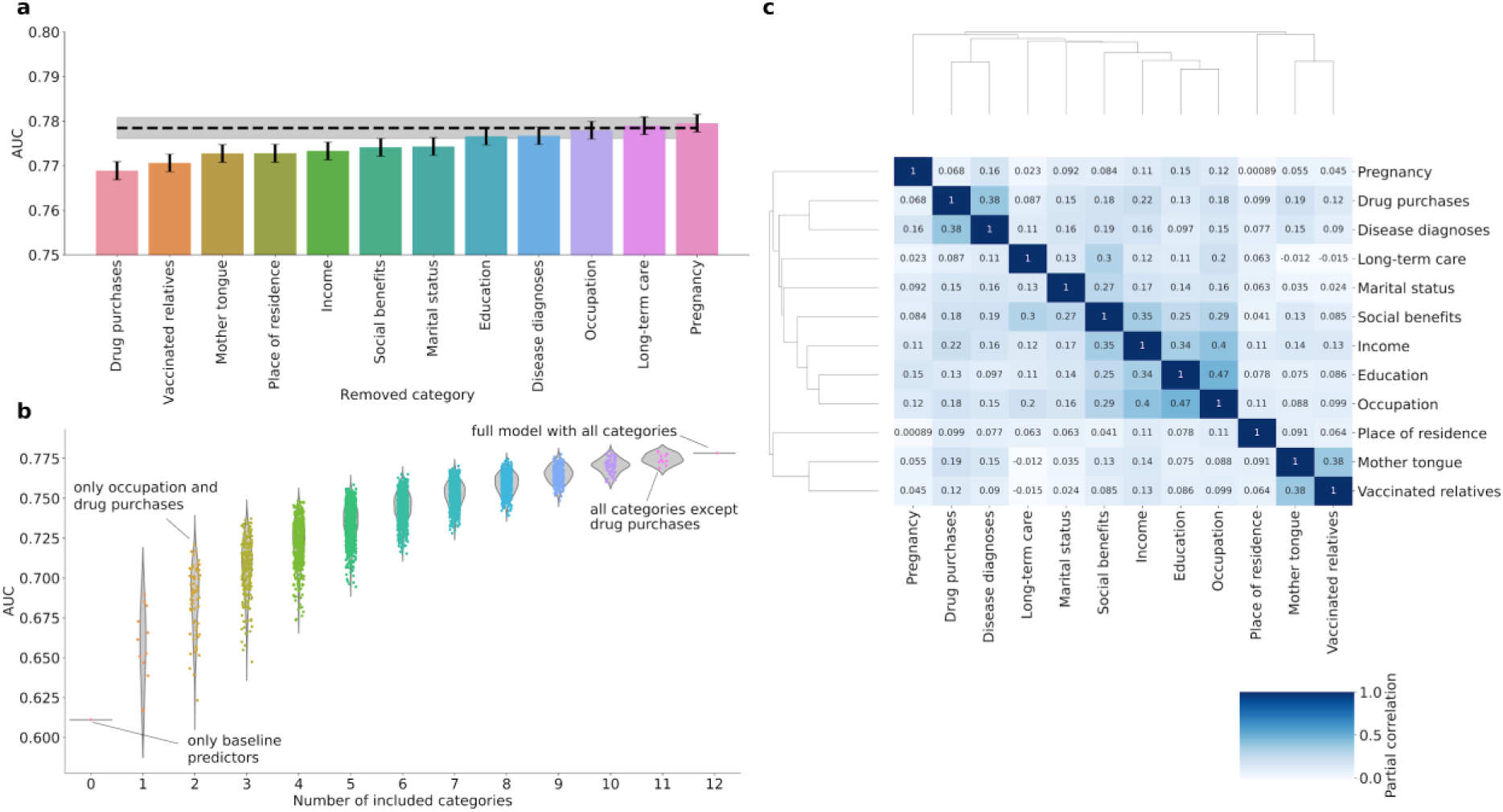
**a)** Drop in AUC (area under receiver-operator characteristics curve, y-axis) when removing a single category at a time from the full Lasso classifier (including all predictors). Removing all predictors from a category removes all information unique to the predictors of that category, meaning that the drop in AUC quantifies the loss in predictive power due to information unique to the removed category. The lower the AUC here, the higher is the amount of unique information contained in the category that is useful for predicting COVID-19 vaccination uptake. The black dashed line indicates AUC of the full Lasso model using all predictor categories. Error bars and the error band correspond to 95% confidence intervals computed using bootstrapping. **b)** Drop in AUC (area under receiver-operator characteristics curve, y-axis) when removing different combinations of predictor categories from the full Lasso model (full model corresponds to “Number of included categories = 12”). All combinations of removed categories were tested by training separate Lasso classifiers on the data including only the specific combination of predictor categories, and the corresponding AUCs are shown as individual dots. Violin plots show the distribution of AUCs for each number of removed categories. Individual models discussed in the text are highlighted and named. The model with 0 removed categories corresponds to a model trained using the baseline predictors age and sex only. All models include also age and sex as predictors. Panel 4a shows a detailed view of “Number of included categories = 11”. **c)** Pairwise partial Pearson correlation, adjusting for age and sex, between predicted probabilities of COVID-19 vaccination uptake for each test set sample, obtained from each category separately (XGBoost classifiers, AUCs for these models shown in **Fig. 2a** and **Supplementary Table 1**). Color indicates the strength of correlation, and the correlation coefficient is shown on each heatmap cell. Hierarchical clustering dendrograms of the partial correlation matrix of model predictions are shown beside the matrix and were used in ordering the rows and columns.

We then proceeded to study the impact of removing multiple categories simultaneously on the prediction of COVID-19 vaccination uptake. This allows us to identify category combinations that have the largest effect on the model predictions (**Figure 4b**). For example, removing 10 out of 12 categories results in only 7.4% AUC decrease (with the most predictive model containing two categories, occupation and drug purchases) compared to the full model with all 12 categories. Taken together, these results indicate that predictive information is substantially shared across categories and relatively good prediction accuracy can be achieved even in settings where some of the information used in this study are missing.

To understand how much predictive information was shared across categories, independently of age and sex, we computed partial pairwise Pearson correlation between predicted probabilities obtained from models trained separately in each category (**Figure 4c**, same models shown in **Figure 2a**). We found that COVID-19 vaccination uptake probabilities predicted using income, education, occupation and social benefits categories were highly correlated and clustered together (Pearson partial correlation coefficient > 0.25). We also identified significant correlations between predicted probabilities from socio-economic categories and health-related categories. For example, the correlation between predicted probabilities from income and drug purchase history categories was 0.22.

### Genetic information is a weak predictor of COVID-19 vaccination, but correlates with COVID-19 severity and behavioral traits

We performed a genome-wide association study of COVID-19 vaccination uptake in FinnGen (N=273,615) and the Estonian Biobank (N=145,615), restricted to European ancestry. Effects were consistent across the two studies as evidenced by a genetic correlation of 0.8 (95% CI: 0.66-0.95). We, therefore, performed a meta-analysis using METAL [20]. We identified 8 genome-wide significant loci (P-value ≤ 5×10^−8^) (**Figure 5a; Methods**) and, in **Supplementary Table 5**, we reported the most likely gene linked to each lead variant by using a machine learning-based prioritization score from Open Targets Genetics [21,22]. Four out of eight lead variants were associated with anthropometric traits, such as body fat distribution (**Supplementary Table 5**). These four variants increased the likelihood of vaccination while being associated with reduced body fat. We next investigated the SNP-based heritability of vaccination uptake through LDSC regression [23], finding a low but significant SNP-based heritability (observed scale h^2^_SNP_ = 2.6%, SE = 0.18%, p-val = 1.36×10^−47^).

**Figure 5.**
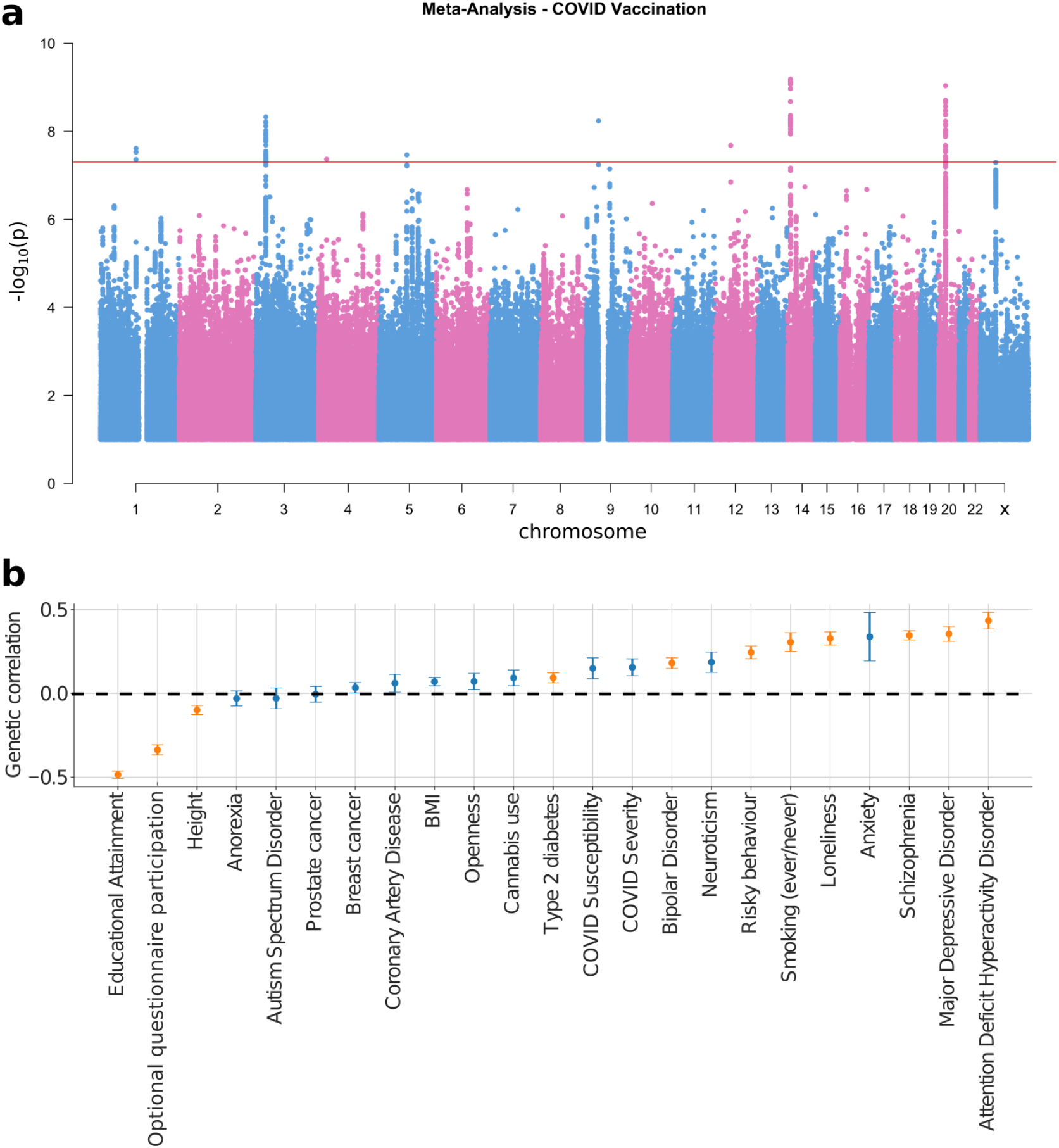
**a)** Manhattan plot of COVID-19 vaccination uptake from meta-analysis of FinnGen and Estonian Biobank. Genetic variant must have been tested in both datasets, passed quality control in both (INFO ≥ 0.8 and MAF ≥ 0.1%) and significant variants must not have indicated significant heterogeneity (heterogeneity P-value < 0.0056 - P-value corrected for multiple testing with 9 significant variants). The red horizontal line indicates genome-wide significance. **b)** Genetic correlations between COVID-19 vaccination uptake and selected health and behavioral phenotypes. Error bars represent standard errors. Orange error bars and point estimates represent Bonferroni-significant genetic correlations (P-value < 0.002). The black dashed line indicates 0 genetic correlation. Positive correlation means correlation with reduced COVID-19 vaccination uptake. BMI = Body Mass Index.

Given the significant heritability, we explored if we could build a polygenic score (PGS) that is predictive of vaccination uptake. We re-ran the GWAS on 70% of the FinnGen individuals, meta-analyzed these results with the GWAS conducted in the Estonia Biobank, and used the results to build a PGS in the remaining 30% of the FinnGen individuals. A model including age, sex and the PGS reached an AUC of 0.612 (95% CI: 0.601 - 0.623) when predicting vaccination uptake, significantly higher than the baseline model including only age and sex (AUC=0.589, 95% CI: 0.578 - 0.600, P-value for improvement=1.72×10^−9^). PGS predicted vaccination status better than the pregnancy and long-term care categories, and similarly to municipality of residence (**Supplementary Table 1**).

We explored the genetic correlations between the GWAS of vaccination uptake and a series of other health and behavioral information, mostly not available in the nationwide FinRegistry dataset. Of the 23 phenotypes tested, 11 were significant after multiple hypothesis testing correction (P-value < 2 × 10^−3^ (Bonferroni correction for 23 tests)) (**Figure 5b**). Four psychiatric disorders - schizophrenia, major depressive disorder, bipolar disorder and attention deficit hyperactivity disorder - were all positively genetically associated with reduced vaccination uptake (*rg* between 0.18 and 0.43), consistent with the epidemiological results (**Figure 2c)**. Not vaccinating was also associated with a higher genetic predisposition to loneliness, risky behavior and smoking (*rg* between 0.25 and 0.33). Interestingly, we found a negative correlation (*rg*= -0.34, 95% CI: -0.40 - -0.28) with participation in subsequent questionnaires of UK Biobank (a proxy for engagement in scientific research) (**Supplementary Table 6**). Genetic correlations were comparable when COVID-19 cases were included in the vaccination uptake phenotype (**Extended Data Figure 6;** only for FinnGen study).

To test if individuals at higher genetic risk for COVID-19 susceptibility and severity were more or less likely to vaccinate, we built PGS for COVID-19 severity and susceptibility using Release 7 from the COVID-19 host Genetic initiative [15], which includes mostly studies collected before the start of the vaccination campaigns. Individuals with higher PGS for COVID-19 severity and susceptibility were less likely to receive the vaccine. However, the association was modest (severity: OR = 1.03, 95%CI: 1.02 - 1.05; susceptibility: OR = 1.02, 95%CI: 1.01 - 1.04 per 1 standard deviation in PGS) partially due to the PGS for COVID-19 being a weak predictor of COVID-19. Mendelian randomization analysis indicates a lack of causal relationship between COVID-19 severity and reduced vaccination uptake (**Supplementary Table 9**).

## Discussion

The digitalization, harmonization, and accessibility of information routinely collected within healthcare and by governmental agencies opens the possibility to inform policymakers at an unprecedented pace and breadth. The comprehensive collection of nationwide registers combined with biobank data and empowered by machine learning approaches allowed us to extensively compare the impact that health-related, socioeconomic, familial, genetics and demographic information have on one the most pressing public health issues: participation in COVID-19 vaccination programs.

Even in the relatively equal Finnish society, socio-economic aspects and, in particular, labor income in 2019, or lack thereof, were the strongest predictors of uptaking the first dose of COVID-19 vaccine. This observation could also be partly explained by people in lower income occupations having more limited access to vaccines due to e.g. stricter working schedules. Nonetheless, information about job professions was a weaker predictor of vaccination uptake than income. Lack of income in 2019, the strongest predictor, captures a wide range of socio-economic factors including unemployment, severe illness, and retirement.

Several disease conditions were associated with vaccination uptake. Mental health was the most important category: especially psychosis-related conditions and diagnoses related to substance use disorders were associated with lower vaccination uptake. Associations with individual drug purchases supported these observations. People with mental health disorders are at increased risk of (severe) COVID-19, but even more notably, COVID-19 can cause deterioration in mental health, reduction in neuropsychiatric functioning, and even neurodegeneration [24]. As those suffering from mental health disorders appear to be at higher risk of non-vaccination, efforts to increase their vaccination uptake could prove especially effective in reducing both the acute infections and the multifactorial burden of long COVID at an individual and societal level.

Interestingly, drugs used in the management of Alzheimer’s and Parkinson’s diseases were associated with lower vaccination rates. People with these conditions are at higher risk for severe disease from COVID-19 [25], likely have reduced functioning in everyday life and can also have reduced ability to make informed decisions about their vaccination. Other, more common diseases were associated with reduced vaccination uptake, likely by capturing underlying socio-economic factors.

Previous studies have shown that the experience of a family member with COVID-19 increased acceptance of the COVID-19 vaccine [26]. In line with this observation, we found that vaccination status correlates within families. For example, having an unvaccinated mother increased the risk of not being vaccinated (OR=1.31, 95% CI:1.31-1.32). The observation that family members influence vaccination uptake with different magnitudes indicates that other factors, beyond those shared within families (e.g. socio-economic status), impact vaccination status.

History of medication purchases was the strongest predictor category together with income, despite none of the individual drugs being a very strong predictor alone. We hypothesize that patterns of drug purchases are a relatively good proxy for both health and socio-economic aspects. In light of this observation, we performed extensive analyses to understand if different predictor categories are capturing overlapping information. We found a large overlap and redundancy in the predictive properties of different categories, some of which are traditionally considered independently (e.g. health and socio-economic indicators). This observation is important for two reasons. First, it blurs the distinction between health and socio-economic information. Contamination between these two categories has consequences for law and ethics scholars. For example, informed consents in biomedical studies are often bound to health-related research, while we show that socio-economic and health information can capture similar underlying aspects in predicting vaccination uptake. Second, it questions the feasibility of excluding information perceived as sensitive from machine learning-based prediction models. For example, citizens might be against using income to identify individuals at higher risk of not vaccinating but be more inclined to accept targeting individuals based on certain previous health conditions. We showed that predicted probabilities of vaccination uptake obtained using drug purchase history are correlated with predictions obtained using income, questioning whether drug purchase information should be used in a hypothetical scenario where income cannot be used as a predictor.

We showed that by including all the ∼3,000 predictors we could train a well-predictive model of COVID-19 vaccination uptake. For example, such a model can be used to identify 1% of the population with an average vaccination rate of approximately 19%, which is almost 5 times lower than the national average. A simpler prediction model can likely be constructed without a large loss of predictive power, using few socio-economic variables.

GWASs have been conducted on thousands of health and behavioral traits and many behavioral traits are affected by genetic factors [27]. Importantly, both COVID-19 susceptibility and severity have an important genetic component [15]. This, together with the observation that many of the registry-based predictors identified in this study have been shown to have a strong genetic risk component (e.g. mental health disorders) led us to study genetic predictors of COVID-19 vaccination uptake. Genetic information is measured from many individuals (up to 10% of the Finnish population by 2023), has low measurement error, is stable through life, and is not impacted by reverse causation. For the above-listed reasons, statistical genetics approaches can be used to identify correlates of vaccination uptake that are not easily measured nationwide. Such correlates can be easily tested for replication in datasets from different countries. We demonstrate this by performing a meta-analysis on FinnGen and Estonian Biobank study participants showing that genetic correlations between vaccination uptake and socioeconomic traits or psychiatric disorders persist across countries. Interestingly, we found a significant genetic correlation with participating in optional questionnaires within the UK Biobank, supporting a shared underlying effect between participating in scientific studies and propensity to vaccination. We also found that genetic information summarized in PGS are weak, but not insignificant, predictors of COVID-19 vaccination. Finally, our results indicate that individuals at higher genetic risk of severe COVID-19 are less likely to get vaccinated but that this association is unlikely causal and more likely due to shared risk factors captured by the PGS.

Our approach has several limitations. First, generalizability outside Finland and to non-European ancestries is unclear and replication in other countries is needed to understand the generalizability of our findings across different populations. Previous studies using nationwide registers have, however, shown similar risk factors for severe COVID-19 as in other countries [28,29]. Second, information about deaths and emigration from Finland during the year 2021 was not available to us. Thus, some individuals might not have taken the COVID-19 vaccination because they had passed away or had emigrated during the follow-up period. We restricted the analyses to individuals younger than 80 years old to reduce the number of individuals expected to die in 2021. Third, due to the scope and complexity of the included predictors, we made some simplifying decisions in preprocessing the nationwide registry data. Thus, the predictors included in the analyses are subject to some simplifications and limitations. We considered disease diagnoses and drug purchases over the lifespan of the individuals in the study population and condensed this information into binary yes/no predictors. Missing values for many socio-economic variables were considered by including separate predictors for missingness, but there might be multiple reasons for missing records. Better modeling of missing data and age of diagnosis are likely to further increase the predictive performance of the models presented in this study. Not everyone reports emigrating outside Finland to the responsible authorities. To capture this potential bias, we performed a sensitivity analysis removing individuals with no data entries in the year 2019 and showed, overall, no significant changes in the AUCs of individual predictors.

In conclusion, by performing a rapid nationwide examination of predictors of COVID-19 vaccination uptake across different life domains, we have highlighted the importance of harmonized and accessible registry and biobank-based information. We have shown that COVID-19 vaccination uptake is multifactorial and that individuals at higher risk of suffering the worst consequences of COVID-19 are also those less likely to uptake COVID-19 vaccination. These results provide potential avenues for targeted interventions supporting COVID-19 and possibly other national immunization programs.

## Data and Methods

### Study population

The FinRegistry dataset (https://www.finregistry.fi/), used in the phenotypic analyses, includes 7,166,416 individuals of whom 5,339,804 (74.51%) are index individuals (every resident of Finland alive on January 1st 2010). The remaining 1,826,612 individuals are relatives (offspring, parents, siblings) and spouses of index individuals who are not index individuals themselves.

FinnGen [16] and the Estonian Biobank (EstBB) [30] were used to explore the role of genetics in COVID-19 vaccination status. The FinnGen project combines multiple hospital biobanks and digital health registries. The data release used for this analysis (Release 9) has genotype data available for 377,277 individuals of Finnish ancestry. The Estonian Biobank is a volunteer-based sample with continually updated national health registry linkage and genotype data on 202,910 individuals.

To restrict the study population to individuals who had had a fair opportunity of receiving the first dose of a COVID-19 vaccination by the end of October 2021, we excluded the following individuals:

1. Individuals who had died or emigrated before 31.12.2020 (death statistics for year 2021 in Finland were not available).
2. Individuals who were less than 30 years old at 31.10.2021.
3. Individuals who were older than 80 years old at 31.10.2021.
4. Individuals who had a laboratory-confirmed COVID-19 diagnosis prior to 31.10.2021.
5. Individuals living in a municipality called Askola.

For the genetic analyses conducted in FinnGen and Estonian Biobank, death or emigration was limited to 31.12.2019 as statistics beyond this date were unavailable for FinnGen. Residents of Askola were excluded, as it was the only municipality where the vaccination coverage differed radically from any other Finnish municipality (see **Extended Data Figure 1**). After these exclusion criteria, the final FinRegistry study population contains 3,192,505 individuals, the FinnGen study population includes 273,615 individuals.

The study outcome, having received at least one dose of a COVID-19 vaccine by 31.10.2021 was defined for the Finnish data using the official registry-based definition by the National Institute for Health and Welfare:

1. Identifying all participants with a record for the ATC code J07BX03 (covid vaccinations).
2. Identifying all participants with a record corresponding to a relevant drug name. The criteria included all records with a drug definition or trade name including: “COM”, “COV”, “CVID”, “CO19”,”COR”,”KOR”,”PFI”,”MOD”,”AST”,”AZ”,”BION”,”SPIKE”. From the set of records identified using these criteria, we excluded ambiguous records containing: “TIC”, “ZOSTA”, “NEULA”, “VESIROK”, “DUKORAL”, “TUHKA”, “COVAC”, “VAZ”, “ZAST”, “PASTEUR”, “FLUR”, “LASTEN”, ““KURKKU”, “SUSTA”.
3. Records were only considered after 01.10.2020.

In the Estonian Biobank, the study outcome of having received at least one dose of a COVID-19 vaccine was defined based on linked data from the national Health and Welfare Information Systems Centre (TEHIK). Health care providers in Estonia have to submit all vaccination notifications to TEHIK, which is also the institution responsible for creating vaccination certificates. The database contains the following information: name of vaccine, anatomical therapeutic chemical (ATC) code, amount (mcg), dosing and schedule. We included all individuals with at least one record of a COVID-19 vaccine (ATC code J07BX03) between 10 October 2020 and 31 October 2021 as cases, and others as controls.

### Selection and definition of the phenotypic predictors

The FinRegistry study contains a comprehensive selection of data modalities ranging from disease history to drug purchase history and detailed socioeconomic variables, as illustrated in **Figure 1a**. We performed an initial variable selection by manually curating variables of interest across the different registries. Categorical variables were dichotomized into indicator variables. Individual predictors, and their manually curated categories are listed in **Supplementary Tables 2-3**. For each predictor, excluding disease occurrences and drug purchases, we also included a binary predictor indicating if the value for this predictor was missing or not. For disease diagnoses and drug purchases, not having a record of the diagnosis/purchase was interpreted as absence of the diagnoses/purchase. Taken together, we defined in total 2,997 predictors (including age and sex). Prevalence of the predictors within the study population was not assessed beforehand. To preserve the privacy of individuals in the study population, FinRegistry has a policy that allows exporting aggregated data only when the aggregated data is based on 5 or more individuals. Some of the very rare predictors had fewer than this number of individuals either among vaccinated or unvaccinated, and thus individual predictor level results for these predictors could not be exported from the secure analysis environment. In total, 105 of the defined predictors were excluded from the individual predictor level results due to this, leaving us with 2,892 predictors (including age and sex). Preprocessing of different categories of phenotypic predictors is discussed in more detail below, each category at a time.

#### Drug purchases

Information about drug purchases was retrieved from the Social Insurance Institution of Finland, Kela, which is a government agency that provides basic economic security by financial support for Finnish residents and many Finns living abroad. One of the social security benefits provided by Kela is reimbursements of part of the costs of medicines that are prescribed for the treatment of an illness. This data contains nation-wide information about prescribed drugs that are purchased from pharmacies. It does not include drugs delivered in hospitals or purchases of drugs without a prescription. This register exists from 1995. Drug purchase information was coded into binary predictors describing whether an individual has ever purchased the drug during 1995-2019. Similar drugs were collapsed into one predictor by considering only the first five digits of the ATC-codes.

#### Occupation

Information about job occupation was retrieved from Statistics Finland, which is a Finnish public authority that collects, combines, and stores data on a wide range of topics. Occupation is available for employed people at the end of the statistical reference year. The information exists from years 1970, 1975, 1980, 1990, 1993, 1995, 2000, and 2004 on an annual basis. We defined occupation as the latest reported (not unknown) occupation before 31.12.2019. Occupation information was coded into 11 binary predictors, according to the highest-level categorization in the Statistics Finland data.

#### Disease history

Disease history was captured using two sets of data, FinnGen clinical endpoints and Finnish National Infectious Diseases Register. The clinical endpoints have been originally defined for the FinnGen project [16] by a group of clinical experts. The clinical endpoints were predominantly generated by combining ICD8, ICD9 and ICD10 codes retrieved from the Finnish Institute of Health and Welfare registries (hospital discharge, cause of death, and cancer registers). In addition, for a small proportion of clinical endpoints, information about drug purchases (Kela), drug reimbursements (Kela), surgical procedures (Finnish Institute of Health and Welfare), and primary health care ICD codes (Finnish Institute of Health and Welfare) were utilized. Clinical endpoints were filtered by excluding endpoints with less than 1,000 individuals in the FinRegistry population, and redundant and highly correlated clinical endpoints as defined by FinnGen. Clinical endpoints defined solely based on ATC codes were also excluded as they capture the same information as drug purchases. For more information about FinnGen clinical endpoints and their definitions see https://www.finngen.fi/sites/default/files/inline-files/FinnGen_Endpoints_Elisa%20Lahtela.pdf and https://risteys.finregistry.fi/. Clinical endpoints were collected between 1.1.1969 and 31.12.2019.

The Finnish National Infectious Diseases Register, retrieved from Finnish Institute of Health and Welfare, is based on the Communicable Diseases Act and Decree that requires medical doctors and laboratories to report cases of certain infectious diseases. The data exists from years 1995-2021. The 10 most frequently reported infectious diseases were included as binary variables (having ever had the diagnosis, prior to 31.12.2019), excluding COVID-19. COVID-19 diagnoses (up until the end of the study period 31.10.2021) from the infectious diseases register were used to exclude people from the study population, as people with a COVID-19 diagnosis had different eligibility criteria for vaccination as the rest of the population. In total, the Diseases category includes 1,959 binary predictors that describe if the individual has ever had the diagnosis.

#### Income

Information about income was retrieved from Finnish pension registry. The income covers salary from labor, not income from benefits or capital income. Income from the year 2019 was used as a continuous predictor. Year 2019 was selected as it was the latest full year before the outbreak of the COVID-19 pandemic in Finland. Individuals with missing income information from 2019 (N=1,173,047) were treated as missing data and were not included in computing the income percentiles in **Figure 2c**. Missing income information was treated as a separate binary predictor. There are multiple reasons why income information might be missing including unemployment, severe illness, and retirement.

#### Education

Information about education level and field of education were retrieved from Statistics Finland as the highest completed degree by statistical year. The data exist for years 1970, 1975, 1985, and for every year between 1987 and 2018. Education level was defined as the highest completed degree by the end of 2018, and the field of education used was the field corresponding to the highest completed degree. Education level was coded into 10 binary predictors, according to the highest-level categorization in the Statistics Finland data, with the exception of adding one predictor corresponding to possibly ongoing education. Each individual aged between 30-35 was assigned to this category based on the median age of receiving doctoral degree in our dataset. Correspondingly, the field of education was set to “education possibly ongoing” for everyone aged between 30-35. In total, the field of education was coded into 13 binary predictors.

#### Marital status

Information of the marital status in the study population was retrieved from the Finnish Population Registry from the Digital and Population Data Services Agency. The data exists between 1960 and 2019. Marital status was coded into 9 binary predictors using the latest known marital status. In addition, separate predictors of ever being married or ever divorced were defined based on the same original data.

#### Social benefits

The amount and duration of social benefits received were retrieved from the Finnish Register of Social Assistance. This register covers years between 1985 and 2019, and includes social benefits received by social service clients who, due to lack or insufficiency of income or social security benefits, have claimed social assistance. Social security benefits are not included in the “social benefits” category in this study.

The social benefits data used in this study is a combination of recipients of primary social assistance, preventive social assistance and rehabilitative work benefit. The social benefits category includes four predictors: total actual income support in euros received by an individual between 1985-2019, total number of months an individual has received actual income support between that same interval, total number of months an individual has received any income support, as well as whether an individual has ever received social assistance.

#### Long-term care

Care Register for Social Welfare from Finnish Institute of Health and Welfare was used to obtain information about long-term care periods. This register contains data on activities and clients of institutional care and residential services of social welfare, and covers years between 1995 and 2019. The register contains comprehensive data from individuals who have been clients in: private and/or public retirement homes, elderly 24-hour residential accommodation, institutional care and assisted living for the intellectually disabled, 24-hour residential housing for severely physically or intellectually disabled, treatment for substance abuse, and rehabilitation facilities, or non-round-the-clock housing services.

In this study, we used this data to create two sets of binary predictors. The first set contains 20 different predictors that detail the type of care given to an individual (for example living in an elderly home or rehabilitation facility). The second set contains 29 different predictors that describe the main reason for entering the treatment. In addition, a predictor was created describing whether an individual had any periods of long-term care between 1995-2019 and another predictor to sum up the total number of treatment days within the same period.

#### Place of residence

The latest known place of residence was extracted from the Finnish Population Registry (Digital and population data services agency) on a municipality level. All individuals living in municipality Askola were discarded due to the vaccination coverage in Askola being a heavy outlier. Thus, place of residence was encoded as 306 binary predictors, including a predictor describing whether the place of residence is unknown.

#### Mother tongue

Information about mother tongue was obtained from the Finnish Population Registry from the Digital and Population Data Services Agency. This information is available between 1960-2019. Each mother tongue was considered as a separate binary predictor. Additionally, a predictor summarizing all other mother tongues than Finnish and Swedish was created.

#### Pregnancy related

Information about pregnancy related variables was obtained from the Medical Birth Register from Finnish Institute of Health and Welfare. The information is available for all births given in Finland between 1987 and 2019. We selected manually a set of 47 predictors from the Medical Birth Register. It is worth noticing that the pregnancy related information was only used for women who have been pregnant.

#### Vaccinated relatives

Information about COVID-19 vaccination status was obtained by combining the vaccination registry with information about familial relationships within the study population retrieved from the Finnish Population Registry from the Digital and Population Data Services Agency. The familial information is available between 1964-2019.

For each individual in the study population, we created separate binary predictors describing the vaccination status of their mother and father. If the mother/father was not included in the study population, the value of the corresponding predictor was marked as missing. There are several reasons why a person’s relative would not be included in the study population. They can be too young (<30 yo), too old (>80 yo), dead, or emigrated.

Additionally, we created a binary predictor describing the vaccination status of possible siblings of each individual in the study population. The value of this predictor was coded as 0 if the individual had siblings and any of them was vaccinated, as 1 if the individual had siblings and none of them was vaccinated, and as missing if the individual had no siblings or information about possible siblings’ vaccination status was not available.

### Train/test split, imputation of missing values

The study population was divided at random into training and test sets. Training set contains 80% of the study population. Only the training set was used in model training and fitting. Test set was reserved for computing the performance of the models. Classification performance was measured using Area Under ROC-curve (AUC), and uncertainty of the obtained AUC values was estimated using bootstrapping by drawing with replacement 2,000 samples from the test set, and computing the 95% confidence intervals. For speeding up the training of Lasso and XGBoost classifiers, the training set was downsampled to include all of the non-vaccinated (308,594 individuals) and 4 randomly sampled vaccinated individuals per each non-vaccinated.

Each binary predictor category (except for drug purchases and disease diagnoses, as described above) includes a binary predictor that encodes whether the value was missing in the registries. For example, education level is encoded with 9 binary predictors describing the education levels and one binary predictor indicating whether information about education level was missing. In the logistic regression analysis, individuals with missing values were discarded from the analysis. This corresponds to a complete case analysis. The number of missing values is shown for each predictor in **Supplementary Tables 2-3**. In the Lasso analyses, imputation was used to keep the data set sizes constant across the compared predictors. Imputation was conducted by drawing new values for the missing values with replacement from the distribution of the non-missing values of the same predictor, assuming that the values are missing at random. In XGBoost analyses, missing values were input to the algorithm as is, letting XGBoost learn the rules for handling missing values.

### Logistic regression

Logistic regression adjusted for age and sex was used to determine association of each binary predictor with vaccination status (1 = not vaccinated, 0 = vaccinated). For each binary predictor, the following model was fit on the training split of the data using the function bigglm from library biglm (version 0.9.2.1):

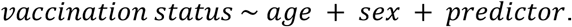

The reference category for the different predictors is detailed in the **Supplementary Table 8**. The p-values of the logistic regression model coefficients were corrected for multiple hypothesis testing using the Benjamini-Hochberg procedure [31], implemented in the Python package statsmodels (version 0.12.2) [32].

### XGBoost classifiers

XGBoost (eXtreme Gradient Boosting, version 1.5.0) [17] classifiers were trained for each predictor category and for the full set of predictors to understand how much learning interactions and non-linearities can boost the vaccination status predictions. All models were trained on the training split of the data using 5-fold cross-validation to optimize the model hyperparameters using Bayesian hyperparameter optimization (BayesSearchCV function from scikit-optimize, version 0.9.0) over the range of possible hyperparameter values detailed in **Supplementary Table 4**, sampling 200 hyperparameter combinations for each model. Balanced class weighting was used.

#### 1) Separate XGBoost classifiers for each predictor category

To determine the predictive performances of the predictor categories, an XGBoost classifier was fitted containing all the predictors from the specific category (see **Supplementary Tables 2-3** for which predictors are included into which category). In addition, age and sex were used as predictors in each model, and a separate baseline model including age and sex only was trained to serve as a benchmark. Results from these XGBoost models are shown in **Figures 2a** and **4c**, and the AUCs are listed in **Supplementary Table 1**.

#### 2) XGBoost classifier trained with the full set of predictors

An XGBoost model was trained using the full set of 2,997 predictors similarly as the individual-category models described above. TreeExplainer-method from the SHAP library [19] was used to interpret the importances of individual predictors of the full XGBoost model in terms of Shapley values. Shapley values were computed averaging over randomly chosen training samples, covering 5% of the whole training set. We used the interventional feature perturbations with a random sample of 100 individuals from the training set as the background data. The results from this model are shown in **Figure 3**. Due to undersampling the vaccinated individuals and using class weights during training, the full XGBoost model is not well calibrated. We used the method proposed in [18] to show that the model can be recalibrated to predict probabilities that correspond well to the actual observed probabilities.

### Lasso classifiers

Lasso classifiers were trained in three slightly different settings: 1) separate Lasso classifiers for each predictor, 2) separate Lasso classifiers for each combination of predictor categories and 3) Lasso classifiers trained with the full set of predictors. All models were trained on the training split of the data using 5-fold cross-validation to optimize the regularization strength. Models were fitted with the cv.glmnet function from the glmnet R package (version 4.1.1) [33] with the default parameter values. Balanced class weighting was used. We separately described the three different settings for training Lasso classifiers in the following.

#### 1) Separate Lasso classifiers for each predictor

To determine the predictive power of individual predictors, a Lasso logistic regression model was fitted for each predictor including age and sex. For each predictor, the following model was fit:

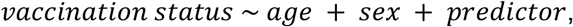

The results from these analyses were used in **Figure 2b**, and the full results are listed in **Supplementary Table 2**.

#### 2) Separate Lasso classifiers for each combination of predictor categories

To quantify the importance of individual predictor categories in forecasting the vaccination status, additional Lasso classifier models were trained with systematically testing each possible combination of the 12 predictor categories. Not including one predictor category in the model removes all information contained only in this predictor from the training data (notice that other predictor categories can partly, or even completely contain the same information that the removed category). For example, excluding all predictors in the Occupation category removes from the training set all information that cannot be explained by any other predictor category. Due to computational complexity of this experiment, requiring training of 4,095 separate models, Lasso was used here instead of the more computationally expensive XGBoost.

To determine the predictive performances of each combination of predictor categories, a Lasso logistic regression model was fitted containing all the predictors from the specific combination of categories. In addition, age and sex were used as predictors in each model. Given a set *C* containing all the predictors in the specific combination, the fitted model is

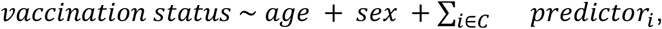

where the index *i* runs over all predictors in category *C*. The results from these analyses were used in **Figures 4a** and **4b**.

#### 3) Lasso classifiers trained with the full set of predictors

To determine the overall predictive performance across all predictors, we trained Lasso logistic regression models also using the full set of 2,997 predictors:

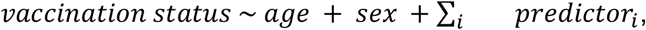

where the index *i* now runs through all predictors. The results from these analyses were used only for comparison with XGBoost, which was chosen as the primary method to calculate the combined prediction model.

### Sensitivity analysis removing individuals with no data entries in the year 2019

As a sensitivity analysis, we removed all individuals with no data entries in the year 2019 and re-run the Lasso and the logistic regression analyses. Specifically, we removed each individual with no disease diagnoses, *and* no drug purchases, *and* no social benefits, *and* no long-term care entries, *and* no birth register entries *and* with zero income. This ended up removing 129,089 out of the total 3,192,505 individuals in the study population, indicating that we have reliable follow-up for a large majority of the study population. We considered only these data sources, because other data sources repeat the entry from the previous year if there is no new entry for the current year. Individuals with no data entries in the year 2019 were removed from the training and test sets, and otherwise the same train/test split was used. The results from this analysis are shown in **Extended Data Figure 4**.

### Calculation of partial correlations between machine learning model predictions, and clustering of predictions

To compute the similarity between the predicted probabilities for COVID-19 vaccination uptake obtained from models trained for each predictor category, we calculated, in the test set, partial Pearson correlations between predicted probabilities from each category and visualized these as a clustered heatmap (**Figure 4c**). To remove the correlation between predicted probabilities which is explained by the fact that age and sex are included in each category, we used the *partial_corr* function from Python library pingouin (version 0.5.2) [34], using default parameters. Clustering of the partial correlation coefficient matrix was computed and the heatmap plotted using the *clustermap* function from Python library seaborn (version 0.11.2) [35], with the default parameters (*method=‘average’, metric=‘euclidean’*).

### Analysis of genetic predictors

We constructed the same vaccination phenotype used for FinRegistry in both FinnGen and Estonia biobank, with the exception that deaths were excluded until 31.12.2019 as data was not available over the full time period (Total: N_cases_=45,202, N_controls_=374,178; FinnGen: N_cases_=19,338, N_controls_=254,427; EstBB: N_cases_=25,864, N_controls_=119,751). GWAS was performed using REGENIE v2.2.4 [36] for FinnGen and SAIGE v1.0.7 [37] for Estonian Biobank (**Supplementary Methods**). To test suitability for meta-analysis, genetic correlations were performed using Linkage Disequilibrium Score Regression (LDSC) and hapmap single nucleotide polymorphisms (SNPs) [23]. Quality control was performed on each set of summary statistics from FinnGen and Estonian Biobank, restricting SNPs to have INFO score ≧ 0.8 and minor allele frequency (MAF) ≧ 0.1%. Meta-analysis was performed using METAL [20]. Genetic correlations with 23 phenotypes - including educational attainment, psychiatric disorders, physical diseases (including COVID susceptibility and severity), anthropometric traits, personality traits and general lifestyle factors - were calculated using Linkage Disequilibrium Score Regression (LDSC) [23] (See **Supplementary Table 3** for a list of summary statistics used for each phenotype).

Polygenic Scores (PGS) for vaccination status were computed using PRS-CS [38]. To remove sample overlap, prior to meta-analysis with the EstBB, we first performed GWAS in a random 70% of the FinnGen study (N_cases_ = 13,555, N_controls_ = 178,081). Association testing was then restricted to the remaining 30%. We trained a logistic regression model of COVID-19 vaccination where the predictors were the vaccination PGS, age and sex by training a regression in 50% of the test set and calculated AUC in the remaining 50%. PGSs for COVID-19 severity and susceptibility were calculated using the same method, but association with COVID-19 vaccination uptake was performed in the full sample due to the lack of sample overlap. COVID-19 Host Genetic Initiative with FinnGen and 23andMe excluded (COVID severity: N_cases_ = 44,549, N_controls_ = 2,018,071; COVID susceptibility: N_cases_ = 155,026, N_controls_ = 2,445,292) were used as summary statistics to calculate PGS [15].

To understand the impact of removing COVID cases on our results, we repeated all analyses including COVID cases within the FinnGen sample.

To test the causal effect of COVID-19 severity on vaccination status, we used Mendelian Randomization [39]. MRBase was used to run two sample mendelian randomization [40]. For the exposure, we selected release 7 of the COVID-19 severity summary statistics with 23andMe and FinnGen samples excluded [15] whereas for the outcome, we selected the summary statistics for vaccination status from the FinnGen sample only as to prevent sample overlap.

## Data Availability

Data dictionaries for FinRegistry are publicly available on the FinRegistry website (www.finregistry.fi/finnish-registry-data). Access to FinRegistry data can be obtained by submitting a data permit application for individual-level data for the Finnish social and health data permit authority Findata (https://asiointi.findata.fi/). The application includes information on the purpose of data use; the requested data, including the variables, definitions for the target and control groups, and external datasets to be combined with FinRegistry data; the dates of the data needed; and a data utilization plan. The requests are evaluated on a case-by-case basis. Once approved, the data are sent to a secure computing environment Kapseli and can be accessed within the European Economic Area (EEA) and within countries with an adequacy decision from the European Commission.
The Finnish biobank data can be accessed through the Fingenious services (https://site.fingenious.fi/en/) managed by FINBB.
Summary statistics of the COVID-19 vaccination uptake GWAS will be made available at the GWAS Catalog upon publication.

## Author contributions

TH, AG, BJ and PV wrote the manuscript with input and comments from all authors. TH performed all analyses using the nation-wide FinRegistry dataset. BJ performed all genetic analyses using FinnGen and all genetic meta-analyses. HS and KK performed the genetic analyses using Estonian Biobank. TH, BJ, AV, PV and AG preprocessed and curated data for the nation-wide FinRegistry analyses. AG, MP, SR, RM, LM and HMO supervised the study.

## Acknowledgements

We would like to thank the entire FinRegistry, FinnGen and Estonia biobank teams for making the data available for the study, and to acknowledge CSC – IT Center for Science, Finland, for computational resources.

This study has received funding from the European Union’s Horizon 2020 research and innovation programme under grant agreement No 101016775. The FinRegistry project has received funding from the European Research Council (ERC) under the European Union’s Horizon 2020 research and innovation program (grant agreement No 945733), starting grant *AI-Prevent*.

This Estonian Biobank study was funded by the European Union through the European Regional Development Fund Project No. 2014-2020.4.01.15-0012 GENTRANSMED. Data analysis was carried out in part in the High-Performance Computing Center of University of Tartu.

We want to acknowledge the participants and investigators of FinnGen study. The FinnGen project is funded by two grants from Business Finland (HUS 4685/31/2016 and UH 4386/31/2016) and the following industry partners: AbbVie Inc., AstraZeneca UK Ltd, Biogen MA Inc., Bristol Myers Squibb (and Celgene Corporation & Celgene International II Sàrl), Genentech Inc., Merck Sharp & Dohme LCC, Pfizer Inc., GlaxoSmithKline Intellectual Property Development Ltd., Sanofi US Services Inc., Maze Therapeutics Inc., Janssen Biotech Inc, Novartis AG, and Boehringer Ingelheim International GmbH. Following biobanks are acknowledged for delivering biobank samples to FinnGen: Auria Biobank (www.auria.fi/biopankki), THL Biobank (www.thl.fi/biobank), Helsinki Biobank (www.helsinginbiopankki.fi), Biobank Borealis of Northern Finland (https://www.ppshp.fi/Tutkimus-ja-opetus/Biopankki/Pages/Biobank-Borealis-briefly-in-English.aspx), Finnish Clinical Biobank Tampere (www.tays.fi/en-US/Research_and_development/Finnish_Clinical_Biobank_Tampere), Biobank of Eastern Finland (www.ita-suomenbiopankki.fi/en), Central Finland Biobank (www.ksshp.fi/fi-FI/Potilaalle/Biopankki), Finnish Red Cross Blood Service Biobank (www.veripalvelu.fi/verenluovutus/biopankkitoiminta), Terveystalo Biobank (www.terveystalo.com/fi/Yritystietoa/Terveystalo-Biopankki/Biopankki/) and Arctic Biobank (https://www.oulu.fi/en/university/faculties-and-units/faculty-medicine/northern-finland-birth-cohorts-and-arctic-biobank). All Finnish Biobanks are members of BBMRI.fi infrastructure (www.bbmri.fi). Finnish Biobank Cooperative -FINBB (https://finbb.fi/) is the coordinator of BBMRI-ERIC operations in Finland.

## Data and code availability

Data dictionaries for FinRegistry are publicly available on the FinRegistry website (www.finregistry.fi/finnish-registry-data). Access to FinRegistry data can be obtained by submitting a data permit application for individual-level data for the Finnish social and health data permit authority Findata (https://asiointi.findata.fi/). The application includes information on the purpose of data use; the requested data, including the variables, definitions for the target and control groups, and external datasets to be combined with FinRegistry data; the dates of the data needed; and a data utilization plan. The requests are evaluated on a case-by-case basis. Once approved, the data are sent to a secure computing environment Kapseli and can be accessed within the European Economic Area (EEA) and within countries with an adequacy decision from the European Commission.

The Finnish biobank data can be accessed through the Fingenious^®^ services (https://site.fingenious.fi/en/) managed by FINBB.

Summary statistics of the COVID-19 vaccination uptake GWAS will be made available at the GWAS Catalog upon publication.

Essential analysis code used to produce the results is available in the FinRegistry GitHub at: https://github.com/dsgelab/COVID-19-vaccination-public.

## Ethics declarations

### Conflicts of interest

None declared.

FinRegistry is a collaboration project of the Finnish Institute for Health and Welfare (THL) and the Data Science Genetic Epidemiology research group at the Institute for Molecular Medicine Finland (FIMM), University of Helsinki. The FinRegistry project has received the following approvals for data access from the National Institute of Health and Welfare (THL/1776/6.02.00/2019 and subsequent amendments), DVV (VRK/5722/2019-2), Finnish Center for Pension (ETK/SUTI 22003) and Statistics Finland (TK-53-1451-19). The FinRegistry project has received IRB approval from the National Institute of Health and Welfare (Kokous 7/2019).

Patients and control subjects in FinnGen provided informed consent for biobank research, based on the Finnish Biobank Act. Alternatively, separate research cohorts, collected prior the Finnish Biobank Act came into effect (in September 2013) and start of FinnGen (August 2017), were collected based on study-specific consents and later transferred to the Finnish biobanks after approval by Fimea (Finnish Medicines Agency), the National Supervisory Authority for Welfare and Health. Recruitment protocols followed the biobank protocols approved by Fimea. The Coordinating Ethics Committee of the Hospital District of Helsinki and Uusimaa (HUS) statement number for the FinnGen study is Nr HUS/990/2017.

The FinnGen study is approved by Finnish Institute for Health and Welfare (permit numbers: THL/2031/6.02.00/2017, THL/1101/5.05.00/2017, THL/341/6.02.00/2018, THL/2222/6.02.00/2018, THL/283/6.02.00/2019, THL/1721/5.05.00/2019 and THL/1524/5.05.00/2020), Digital and population data service agency (permit numbers: VRK43431/2017-3, VRK/6909/2018-3, VRK/4415/2019-3), the Social Insurance Institution (permit numbers: KELA 58/522/2017, KELA 131/522/2018, KELA 70/522/2019, KELA 98/522/2019, KELA 134/522/2019, KELA 138/522/2019, KELA 2/522/2020, KELA 16/522/2020), Findata permit numbers THL/2364/14.02/2020, THL/4055/14.06.00/2020,,THL/3433/14.06.00/2020, THL/4432/14.06/2020, THL/5189/14.06/2020, THL/5894/14.06.00/2020, THL/6619/14.06.00/2020, THL/209/14.06.00/2021, THL/688/14.06.00/2021, THL/1284/14.06.00/2021, THL/1965/14.06.00/2021, THL/5546/14.02.00/2020, THL/2658/14.06.00/2021, THL/4235/14.06.00/202, Statistics Finland (permit numbers: TK-53-1041-17 and TK/143/07.03.00/2020 (earlier TK-53-90-20) TK/1735/07.03.00/2021, TK/3112/07.03.00/2021) and Finnish Registry for Kidney Diseases permission/extract from the meeting minutes on 4^th^ July 2019.

The Biobank Access Decisions for FinnGen samples and data utilized in FinnGen Data Freeze 9 include: THL Biobank BB2017_55, BB2017_111, BB2018_19, BB_2018_34, BB_2018_67, BB2018_71, BB2019_7, BB2019_8, BB2019_26, BB2020_1, Finnish Red Cross Blood Service Biobank 7.12.2017, Helsinki Biobank HUS/359/2017, HUS/248/2020, Auria Biobank AB17-5154 and amendment #1 (August 17 2020), AB20-5926 and amendment #1 (April 23 2020) and it’s modification (Sep 22 2021), Biobank Borealis of Northern Finland_2017_1013, Biobank of Eastern Finland 1186/2018 and amendment 22 § /2020, Finnish Clinical Biobank Tampere MH0004 and amendments (21.02.2020 & 06.10.2020), Central Finland Biobank 1-2017, and Terveystalo Biobank STB 2018001 and amendment 25^th^ Aug 2020.

## Extended Data Figures

**Extended Data Figure 1.**
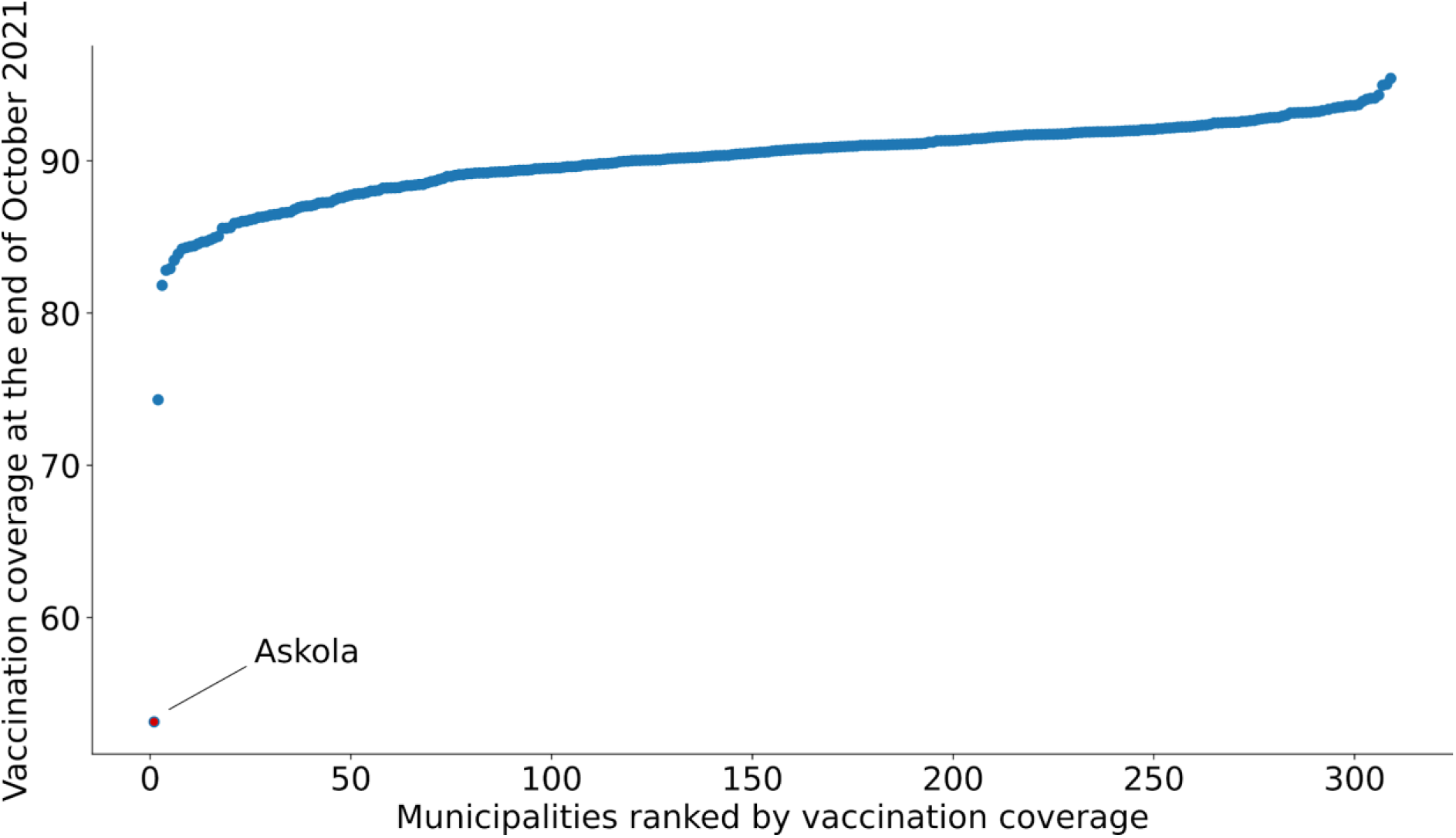
Covid-19 1st dose vaccination coverage in the study population in each Finnish municipality. Residents of Askola (highlighted with red and annotated) were excluded from the study as the vaccination coverage in Askola (2,948 residents in the study population) seemed artificially low compared to all other municipalities and is likely due to misreporting.

**Extended Data Figure 2.**
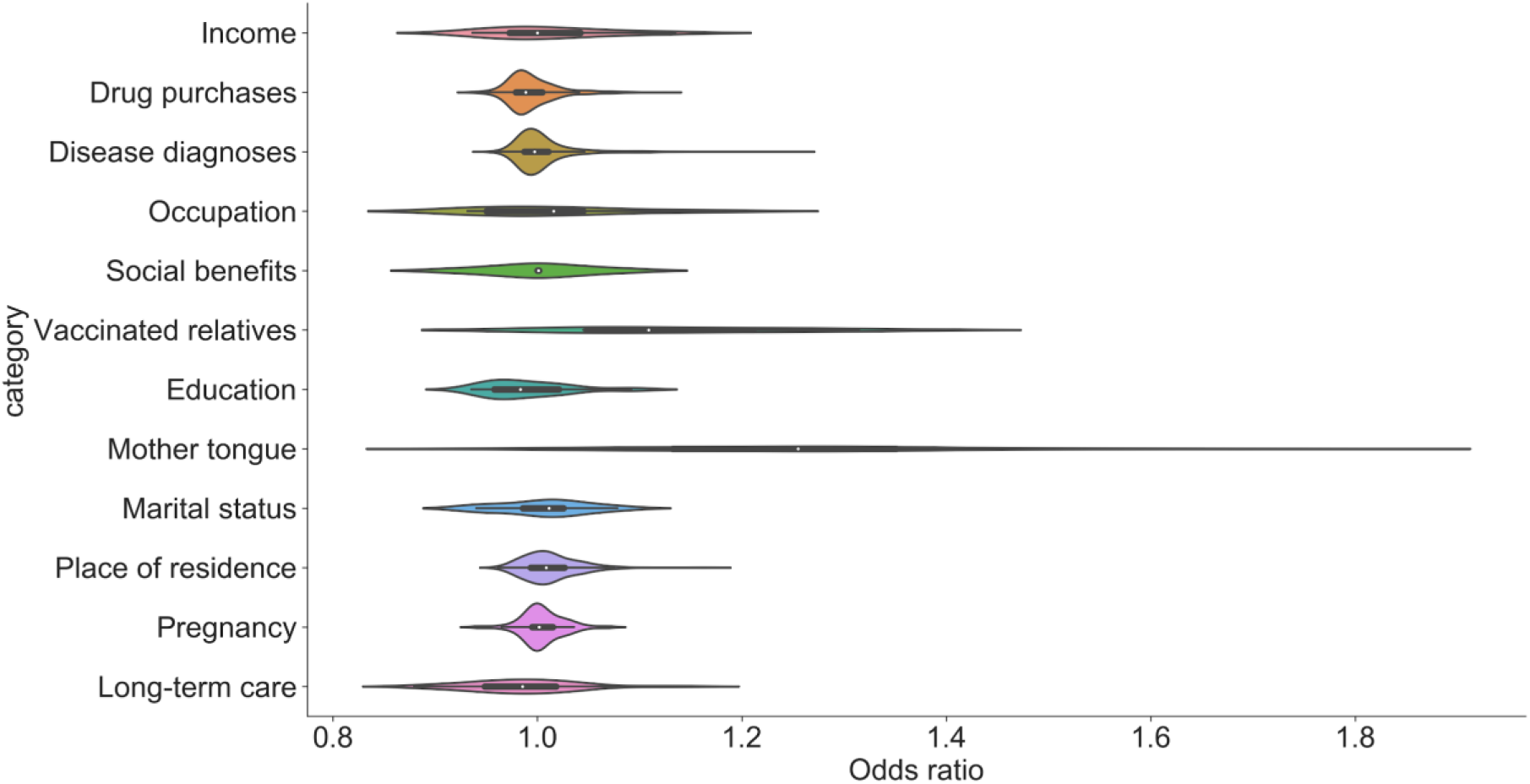
Violin plots describing the distributions of adjusted odds ratios (OR) (adjusted for age and sex, see **Methods**) for not uptaking the COVID-19 vaccination separately for each of the predictor categories. See **Supplementary Table 3** for a full list of ORs for the individual predictors.

**Extended Data Figure 3.**
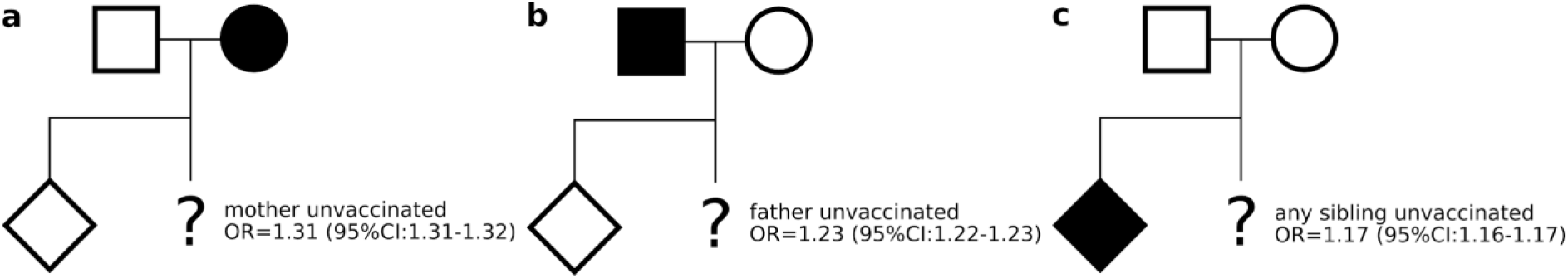
Adjusted (for age and sex, see **Methods**) odds ratios (OR) describing the risk of not uptaking the COVID-19 vaccination when either **a)** mother, **b)** father, or **c)** any of their siblings is unvaccinated (for the entire follow-up period of 1.1.2021-31.10.2021).

**Extended Data Figure 4.**
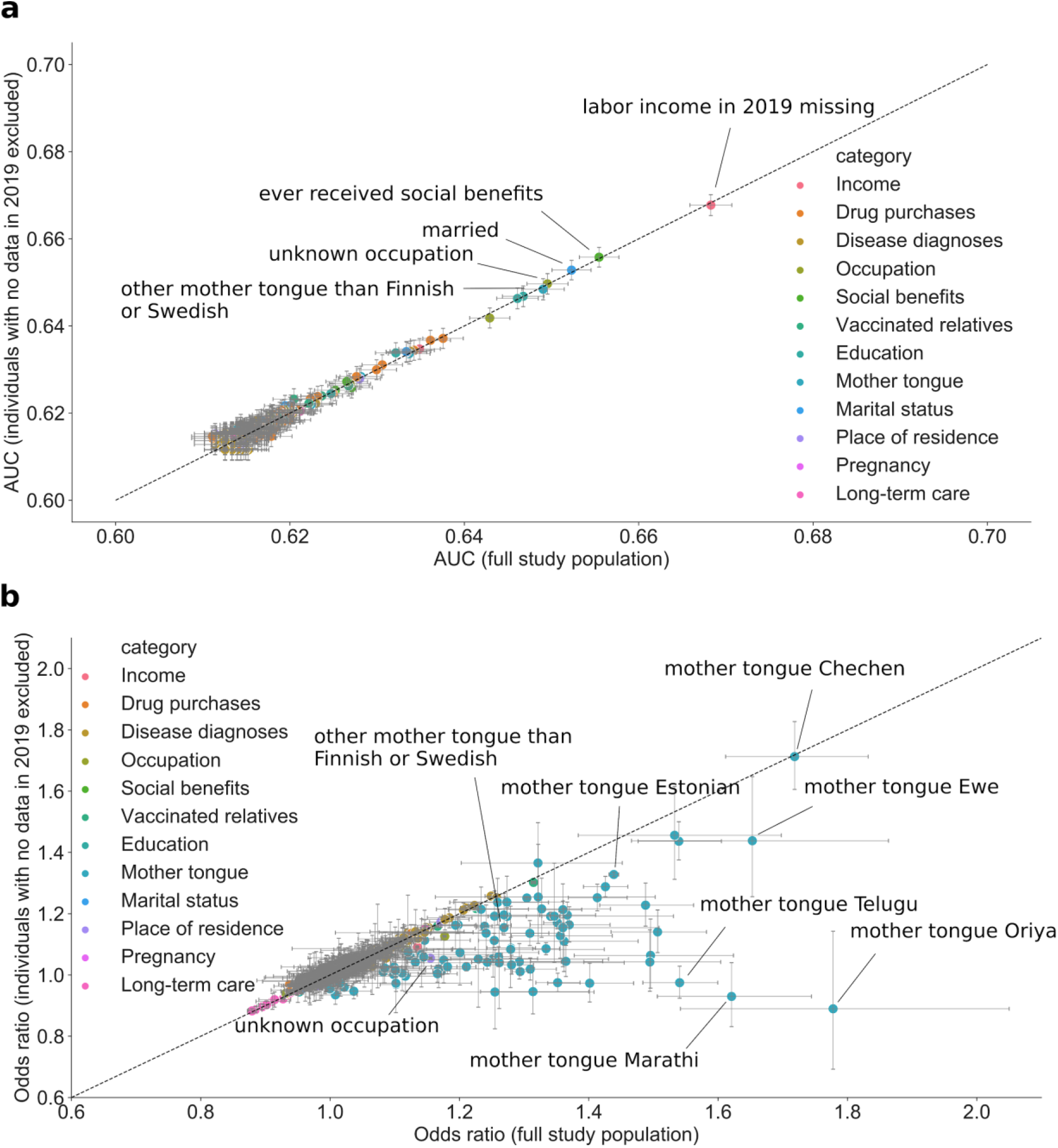
Sensitivity analysis removing all individuals with no data entries in the year 2019 from the study population (in total 129,089 such individuals, see **Data and Methods** for details). The dots are colored by the predictor category. Error bars correspond to 95% confidence intervals computed using bootstrapping. **a)** Area under receiver-operator characteristics curve (AUC) using the full study population (x-axis) plotted against the AUC using the study population with individuals with no data in the year 2019 removed (y-axis) from Lasso classifier models trained separately for each individual predictor (including also age and sex as predictors in each model). Models were trained separately using training data with and without individuals with no data entries in the year 2019. AUCs were computed on a separate unseen test set. No significant changes in AUC were observed for any predictor. **b)** Odds ratios (OR) using the full study population (x-axis) plotted against the ORs using the study population with individuals with no data in the year 2019 removed (y-axis) from logistic regression models trained separately for each individual predictor, adjusting for age and sex. Significant drop in OR when removing individuals with no data in the year 2019 occur mostly for relatively rare mother tongues (some highlighted with labels).

**Extended Data Figure 5.**
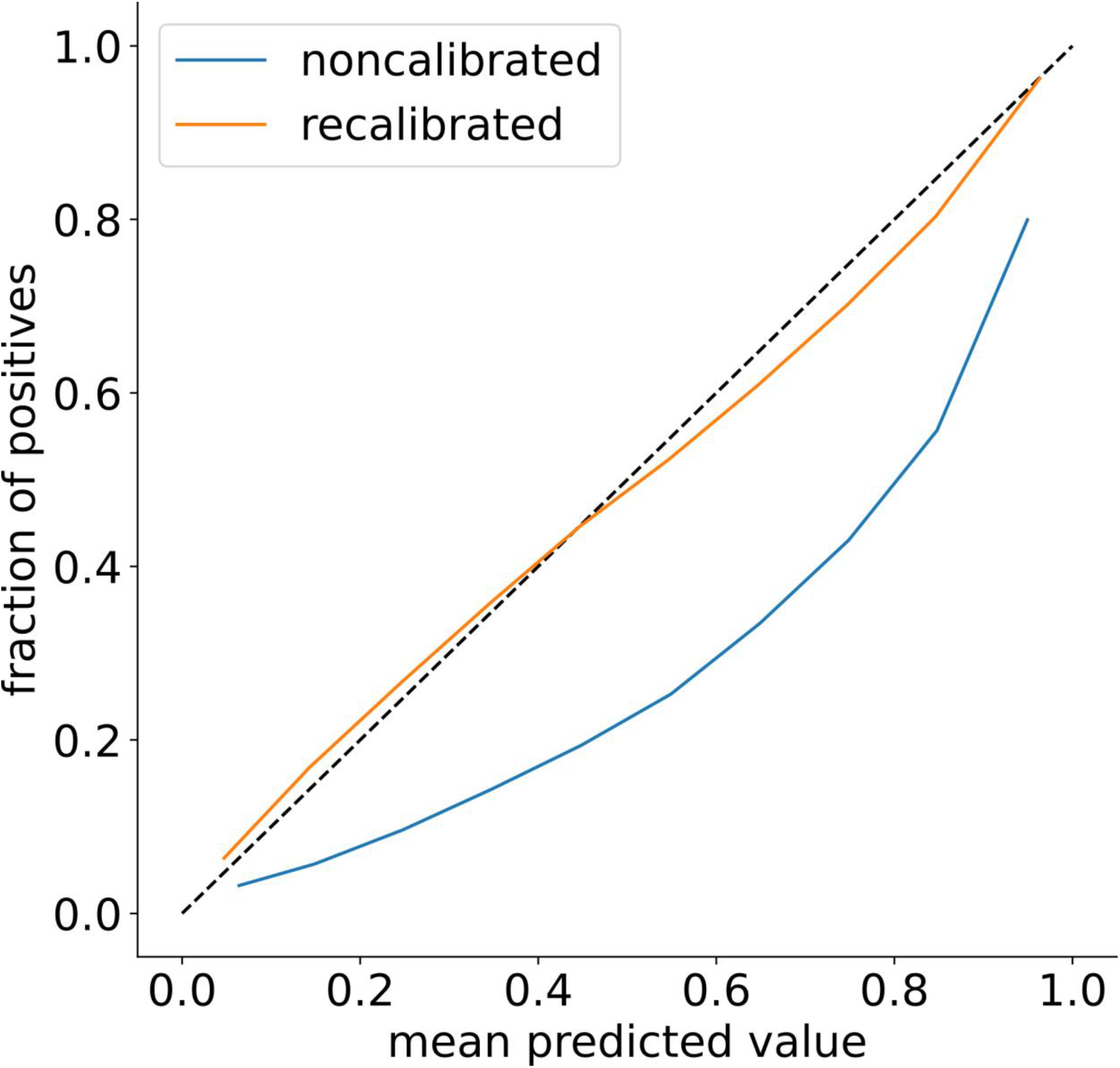
Calibration curves for the full XGBoost (all predictors) model predicting COVID-19 vaccination status before (blue) and after (orange) recalibration (see **Methods**).

**Extended Data Figure 6.**
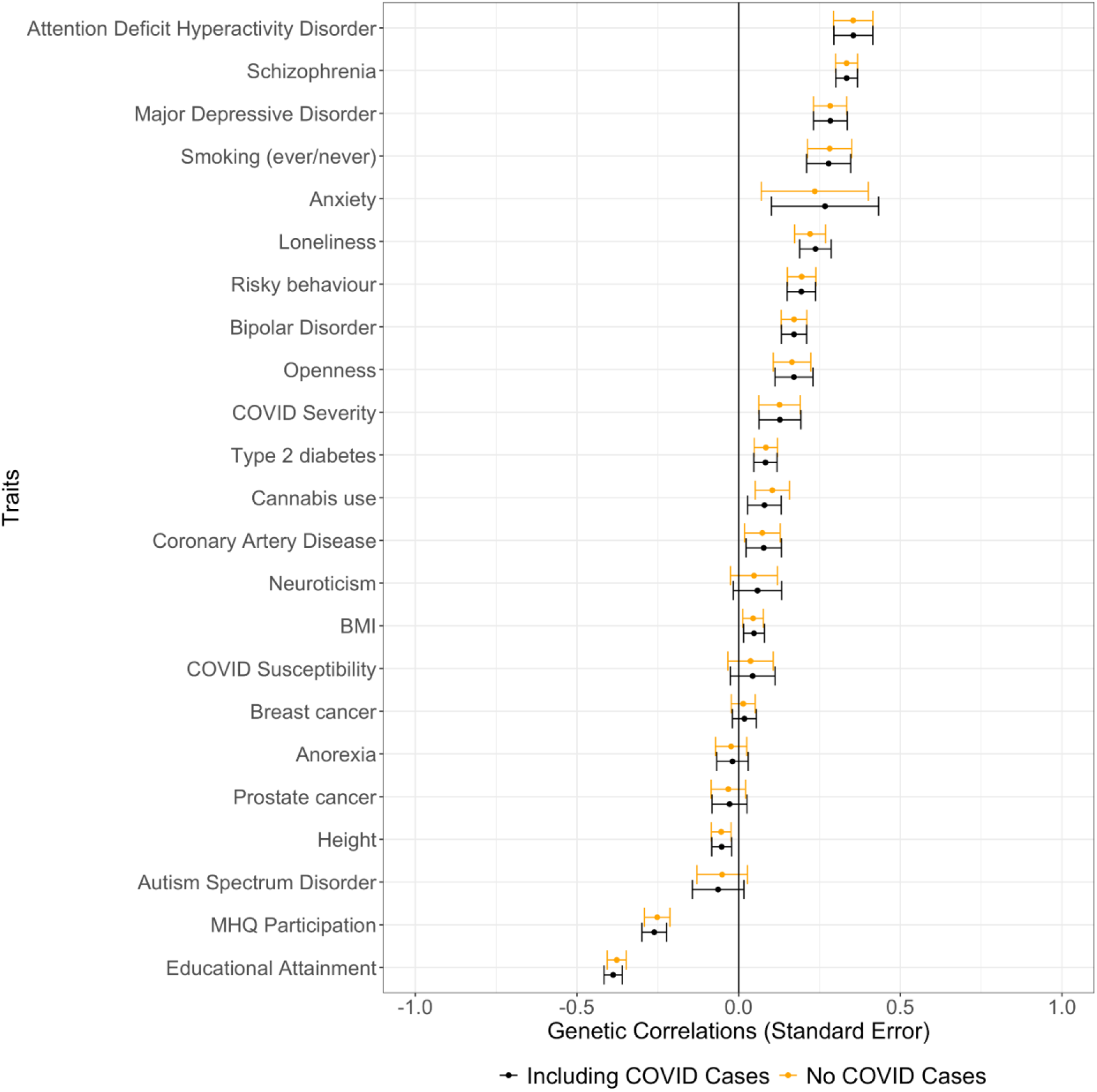
Genetic correlations with and without COVID-19 cases included in the phenotype definition (FinnGen study). Error bars represent standard errors. Black error bars and point estimates represent the vaccination phenotype which includes COVID-19 cases.

## Notes

### Competing Interest Statement

The authors have declared no competing interest.

### Author Declarations

The FinRegistry project has received the following approvals for data access from the National Institute of Health and Welfare (THL/1776/6.02.00/2019 and subsequent amendments), DVV (Digi-, ja vaestotietovirasto) (VRK/5722/2019-2), Finnish Center for Pension (ETK/SUTI 22003) and Statistics Finland (TK-53-1451-19). The FinRegistry project has received IRB approval from the National Institute of Health and Welfare (Kokous 7/2019). Patients and control subjects in FinnGen provided informed consent for biobank research, based on the Finnish Biobank Act. Alternatively, separate research cohorts, collected prior the Finnish Biobank Act came into effect (in September 2013) and start of FinnGen (August 2017), were collected based on study-specific consents and later transferred to the Finnish biobanks after approval by Fimea (Finnish Medicines Agency), the National Supervisory Authority for Welfare and Health. Recruitment protocols followed the biobank protocols approved by Fimea. The Coordinating Ethics Committee of the Hospital District of Helsinki and Uusimaa (HUS) statement number for the FinnGen study is Nr HUS/990/2017. The FinnGen study is approved by Finnish Institute for Health and Welfare (permit numbers: THL/2031/6.02.00/2017, THL/1101/5.05.00/2017, THL/341/6.02.00/2018, THL/2222/6.02.00/2018, THL/283/6.02.00/2019, THL/1721/5.05.00/2019 and THL/1524/5.05.00/2020), Digital and population data service agency (permit numbers: VRK43431/2017-3, VRK/6909/2018-3, VRK/4415/2019-3), the Social Insurance Institution (permit numbers: KELA 58/522/2017, KELA 131/522/2018, KELA 70/522/2019, KELA 98/522/2019, KELA 134/522/2019, KELA 138/522/2019, KELA 2/522/2020, KELA 16/522/2020), Findata permit numbers THL/2364/14.02/2020, THL/4055/14.06.00/2020,,THL/3433/14.06.00/2020, THL/4432/14.06/2020, THL/5189/14.06/2020, THL/5894/14.06.00/2020, THL/6619/14.06.00/2020, THL/209/14.06.00/2021, THL/688/14.06.00/2021, THL/1284/14.06.00/2021, THL/1965/14.06.00/2021, THL/5546/14.02.00/2020, THL/2658/14.06.00/2021, THL/4235/14.06.00/202, Statistics Finland (permit numbers: TK-53-1041-17 and TK/143/07.03.00/2020 (earlier TK-53-90-20) TK/1735/07.03.00/2021, TK/3112/07.03.00/2021) and Finnish Registry for Kidney Diseases permission/extract from the meeting minutes on 4th July 2019. The Biobank Access Decisions for FinnGen samples and data utilized in FinnGen Data Freeze 9 include: THL Biobank BB2017_55, BB2017_111, BB2018_19, BB_2018_34, BB_2018_67, BB2018_71, BB2019_7, BB2019_8, BB2019_26, BB2020_1, Finnish Red Cross Blood Service Biobank 7.12.2017, Helsinki Biobank HUS/359/2017, HUS/248/2020, Auria Biobank AB17-5154 and amendment #1 (August 17 2020), AB20-5926 and amendment #1 (April 23 2020) and its modification (Sep 22 2021), Biobank Borealis of Northern Finland_2017_1013, Biobank of Eastern Finland 1186/2018 and amendment 22/2020, Finnish Clinical Biobank Tampere MH0004 and amendments (21.02.2020 & 06.10.2020), Central Finland Biobank 1-2017, and Terveystalo Biobank STB 2018001 and amendment 25th Aug 2020.

