## Supplementary Information for "Health, socioeconomic and genetic predictors of COVID-19 vaccination uptake: a nationwide machine-learning study"

### Supplementary Methods

***FinnGen***

**Genotyping and quality control**

FinnGen consists of prospectively recruited samples and a series of legacy cohorts with genotypes already available. Prospective samples were genotyped using the ThermoFisher Axiom custom array which tags a total of 655,973 variants. Genotype calling was performed using the Array Power Tools software. Legacy cohorts were genotyped using various Illumina arrays and genotype calling was performed using either GenCall or zCall algorithms.

For both prospective and legacy cohorts the following quality control metrics were used.

Samples were removed if:

- Pihat was > 0.9 and the samples were not monozygotic or replicates
- There was a discrepancy between reported sex and genetically determined sex (F-value ≤ 0.3 for females and ≥ 0.8 for males)
- Missingness was ≥ 5%
- Heterozygosity was ±4 standard deviations from the population average
- Pihat was > 0.1 with 14 or more samples
- Samples were ±4 standard deviations away from the population average according to the first two genetic principal components.

Samples were tagged should there be evidence of a mendelian error or contain replicate samples with over 50,000 discrepancies.

Variants were removed if:

- The variant failed the Hardy-Weinberg Equilibrium test (p-value < $10^{-6}$)
- The variant had a call rate < 98%

**Imputation**

Pre-phasing was performed using Eagle 2.3.5 and samples were imputed using the SiSu v3 imputation reference panel. This reference panel is specific to the Finnish population, containing high-coverage (25-30x) whole-genome sequencing data from 3,775 Finns and 16,962,023 variants with minor allele count ≥ 3. After imputation, 16,387,711 variants were imputed with high quality (INFO > 0.6).

**Ancestry assignment**

Firstly, the FinnGen samples were combined with the 1000 genomes phase 3 dataset. Genetic principal components were calculated using a subset of 49,451 pruned SNPs. Aberrant was used to identify and remove samples that deviated from the main cluster. A probability of belonging to either a North-Western European or Finnish population was calculated by firstly performing PCA with individuals belonging to these ancestries from 1000 genomes data. FinnGen samples were then projected onto this PCA space and Mahalanobis distances calculated for each sample against each of the two ancestries. Samples were retained if there was ≥ 95% probability of belonging to the Finnish ancestry cluster.

***Estonian Biobank***

Estonian Biobank (EstBB) is a population-based cohort with a rich variety of phenotypic and health-related information collected for each participant (Leitsalu et al. 2015). At recruitment, participants signed a consent allowing follow-up linkage of their electronic health records (EHR), thereby providing a longitudinal collection of their phenotypic information. The EstBB database includes health records from the national Health Insurance Fund Treatment Bills (from 2004), Tartu University Hospital (from 2008), and North Estonia Medical Center (from 2005), and data from different registries (causes of death, cancer, etc.). For all the participants EstBB provides information on the diagnoses in ICD-10 coding and information on drug dispensing data, including drug ATC codes, prescription status and purchase date (if available).

##### Genotyping and quality control

Genotyping of DNA samples from the Estonian Biobank was done at the Core Genotyping Lab of the Institute of Genomics, University of Tartu using the Illumina Global Screening Arrays (GSAv1.0, GSAv2.0, and GSAv2.0_EST). Altogether 206,448 samples were genotyped and then PLINK format files were created using Illumina GenomeStudio v2.0.4. During the quality control all individuals with call-rate < 95% or mismatching sex that was defined based on the heterozygosity of X chromosome and sex in the phenotype data, were excluded from the analysis. Variants were filtered by call-rate < 95% and HWE p-value < 1e-4 (autosomal variants only). Variant positions were updated to Genome Reference Consortium Human Build 37 and all variants were changed to be from TOP strand using reference information provided by Dr. Will Rayner from the University of Oxford (https://www.well.ox.ac.uk/~wrayner/strand/). After QC the dataset contained 202,910 samples for imputation.

##### Imputation

Before imputation variants with MAF<1% and Indels were removed. Prephasing was done using the Eagle v2.3 software (Loh et al. 2016) (number of conditioning haplotypes Eagle2 uses when phasing each sample was set to: --Kpbwt=20000) and imputation was carried out using Beagle v.18May20.d20 (S. R. Browning and Browning 2007; B. L. Browning, Zhou, and Browning 2018) with an effective population size ne=20,000. As a reference, Estonian population specific imputation reference of 2297 WGS samples was used (Mitt et al. 2017).

##### Ancestry assignment

Further, EstBB samples were combined with the 1000 genomes phase 3 dataset for ancestry analysis. Genetic principal components were calculated using a subset of quality controlled and pruned genotyped SNPs. This was further used to identify and remove samples that deviated from the main cluster.

### Supplementary Tables

Supplementary Tables are provided in a separate Excel file, here we list just the captions of the tables.

**Supplementary Table 1.** Predictive performance (measured using Area Under Receiver-operator characteristics curve, AUC of the manually curated predictor categories when predicting COVID-19 vaccination status. AUC is reported from XGBoost classifier models (except logistic regression was used for PRS and baseline in FinnGen, see Methods for details) that include all predictors from the corresponding predictor category of interest and age and sex as variables (see Methods for details). Baseline models (including only age and sex as predictors) were trained separately for the population-wide FinRegistry dataset, and the subset of the Finnish population genotyped in FinnGen. Percentage of improved over baseline was computed with respect to the baseline model of the corresponding study population.

**Supplementary Table 2.** Predictive performance (measured using Area Under Receiver-operator characteristics curve, AUC of the individual predictors when predicting COVID-19 vaccination status. AUC is reported from a Lasso classifier model that includes the predictor of interest and age and sex as variables (see Methods for details). If the predictor is not binary, isBinary column value is FALSE. N is the total number of individuals in the study population with the predictor, and N_among_vaccinated the same number among the vaccinated individuals. N_NA describes the number of study population individuals with missing value for the predictor.

**Supplementary Table 3.** Associations between COVID-19 vaccination status and each of the individual predictors. Odds ratios were computed adjusting for age and sex, as described in the Methods. If the predictor is not binary, isBinary column value is FALSE (and N and N_among_vaccinated are missing). N is the total number of individuals in the study population with the predictor, and N_among_vaccinated the same number among the vaccinated individuals. N_NA describes the number of study population individuals with missing value for the predictor.

**Supplementary Table 4.** Hyperparameter spaces tested during training of the XGBoost models. Column names correspond to the XGBoost parameter names. Default values were used for parameters not listed here. Tested parameter ranges are given as Python expressions.

**Supplementary Table 5.** Pleiotropic associations of COVID vaccination lead variants. Discovered using Open Targets Genetics.

**Supplementary Table 6.** Genetic Correlations

**Supplementary Table 7.** Summary statistics used to perform genetic correlations

**Supplementary Table 8.** Reference levels for logistic regression for each predictor categories.

**Supplementary Table 9.** Mendelian Randomization results exploring the causal effect of COVID-19 severity on COVID vaccination
